# Genetic insights into the age-specific biological mechanisms governing human ovarian ageing

**DOI:** 10.1101/2023.03.13.23287201

**Authors:** Sven E. Ojavee, Liza Darrous, Marion Patxot, Kristi Läll, Krista Fischer, Reedik Mägi, Zoltan Kutalik, Matthew R. Robinson

## Abstract

There is currently little evidence that the genetic basis of human phenotype varies significantly across the lifespan. However, time-to-event phenotypes are understudied and can be thought of as reflecting an underlying hazard, which is unlikely to be constant through life when values take a broad range. Here, we find that 74% of 245 genome-wide significant genetic associations with age at natural menopause (ANM) in the UK Biobank show a form of age-specific effect. Nineteen of these replicated discoveries are identified only by our modelling framework, which determines the time-dependency of DNA variant-age-at-onset associations, without a significant multiple-testing burden. Across the range of early to late menopause, we find evidence for significantly different underlying biological pathways, changes in the sign of genetic correlations of ANM to health indicators and outcomes, and differences in inferred causal relationships. We find that DNA damage response processes only act to shape ovarian reserve and depletion for women of early ANM. Genetically mediated delays in ANM were associated with increased relative risk of breast cancer and leiomyoma at all ages, and with high cholesterol and heart failure for late-ANM women. These findings suggest that a better understanding of the age-dependency of genetic risk factor relationships among health indicators and outcomes is achievable through appropriate statistical modelling of large-scale biobank data.

## Introduction

Age-at-onset and time-to-event observations are among the most important traits of interest in cohort studies of age-related diseases, as they are critical to gaining insight into the genetics of disease development and progression [1, 2]. The underlying aetiology of age-related outcomes likely reflects a variety of biological processes that are triggered at different stages of life, long before the onset of observable symptoms. As a result, the underlying genetic propensity for outcomes may vary with age, and depend upon different sets of genetic risk factors at different time points, reflecting the range of underlying molecular mechanisms that shape the onset distribution. Therefore, identifying the genetic variants associated with onset at different stages of life will improve our understanding of disease progression.

Here, we seek to test the hypothesis that genetic propensity for age-at-onset is age-specific, by focusing on the most commonly experienced timing-related phenotype in the human population, age at natural menopause (ANM). Menopause is the permanent cessation of the menstrual cycle in women following the loss of ovarian function and occurs at an average age of 51 years, with 4% of the female population experiencing early menopause prior to age 45. Current evidence suggests that early menopause is associated with a risk for cardiovascular disease [3] and osteoporosis [4], and late menopause is associated with a risk for breast cancer [5]. Recent genomic studies, find ∼ 50% of menopausal timing variation is attributable to genetic markers [6] that are linked to regulation of DNA repair and immune function [7–9]. However, previous analyses make strong assumptions that genetic effects are constant throughout life (Figure 1a). By modelling the quantitative genetic basis of ANM in a way that enables detection of the age at which genetic risk factors have the greatest influence, we report evidence for widespread age-specific genetic effects underlying population-level variation in ovarian ageing in both the UK and Estonian Biobank data.

**Figure 1.**
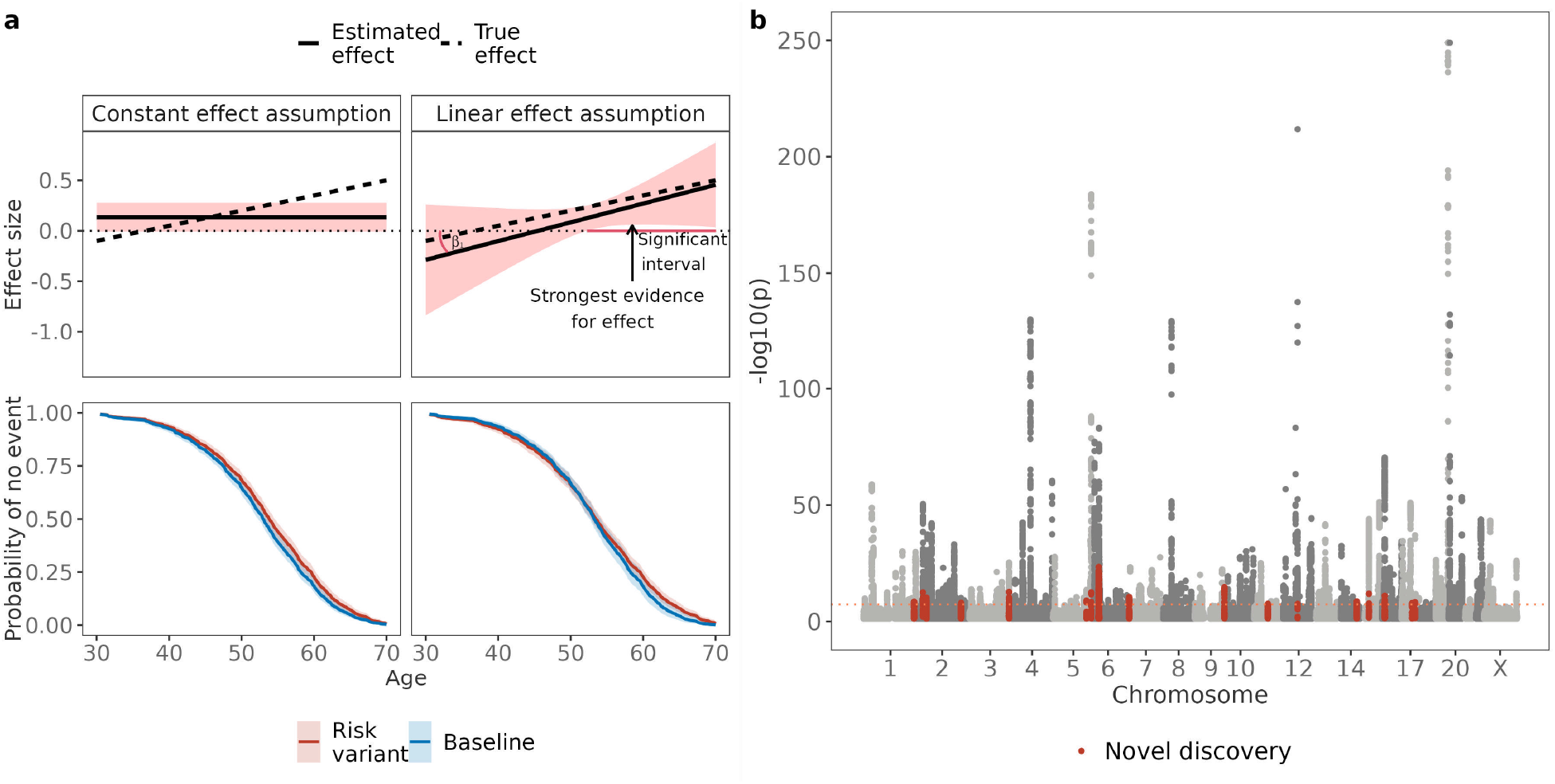
Statistical model description and novel discoveries. (a) The CAMP model enables a more flexible and accurate description of the SNP effect size by introducing a slope term. The linear change model enables three example questions to be addressed: i) what is the interval at which there is a significant effect on the trait? ii) at which age is there strongest evidence for an effect? iii) is the slope (*β*_1_) significantly different from zero? Even though a more complex model can result in generally wider confidence intervals, it can still result in a more accurate representation of the effect size, often accompanied by higher statistical power. By estimating the effect size change it is also possible to accurately determine trends, which constant effect assumptions cannot capture. (b) 19 novel discoveries for age-at-menopause from the CAMP model across the UK and Estonian Biobank data, the coral line indicates the genome-wide significance level of 5 · 10^−8^.

## Results

### Modelling effect size change reveals novel loci

#### Summary of the methods

In the Methods, we present a marginal Cox Age-specific Mixed Proportional hazards model (CAMP) with a novel significance testing framework. CAMP determines the time-dependency of markerage-at-onset associations, without a significant multiple-testing burden. Our two-step approach is a form of parametric Weibull survival analysis, followed by an age-specific Cox proportional hazards model for single marker association testing that can capture the change in genetic hazard for age-at-onset while controlling for the off-locus genetic effects in a mixed-model approximating fashion. Linear-mixed model association testing is widely applied in the genomics field, but here it is explicitly tailored for age-at-onset outcomes with right censoring. In this two-step approach, we first use a BayesW prior [1] to estimate the genetic effects for age-at-onset jointly. That creates the leave-one-chromosome-out genetic predictors that are then used in the age-specific Cox proportional hazards model of the second step. Hence, the final model shape for individual *i*, chromosome *k* and SNP *j* is

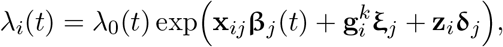

where λ_0_(*t*) is the baseline hazard, **x**_*ij*_ is the standardised genotype for the *j*th marker, 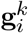 is the BayesW genetic predictor for individual *i* from all chromosomes other than *k*, **ξ**_*j*_ is its corresponding effect when estimating marker *j*, **z**_*i*_ is the summarised covariate value and **δ**_*j*_ is its corresponding effect when estimating marker *j*. 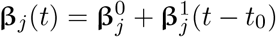 is the effect size change function for SNP *j* which we assume is a linear function (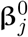 is the intercept, 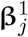 is the slope, *t*_0_ is an offset specifying the intercept interpretation, see Methods). We then test for age-at-onset associations for every SNP given our theoretically supported significance testing procedure (see Methods).

### Discovery of novel loci

We apply our CAMP model to 173,424 unrelated observations of self-reported ANM in the UK Biobank data (125,697 reported events and 47,727 right-censored observations; 8,747,951 SNPs), and 70,082 observations in the Estonian Biobank (22,740 reported events and 47,342 censored observations, Figure S1). We find 312 ANM associations in the UK Biobank, of which 226 replicate previous studies [7–9], and 19 are novel and replicate for the first time within the Estonian Biobank (Table 1, Figure 1b, see Figure S2). In addition, we find 67 associations that have not previously been reported, but they did not replicate in the Estonian Biobank. Nevertheless, 46 out of 67 previously unreported associations show consistency with signs in the discovery and replication data sets (Fisher’s exact test, *p* = 0.007), suggesting that a larger replication data set could lead to further replications. In conclusion, the CAMP approach yields a power increase: 8% of the replicated marker associations are previously unreported, an increase consistent with the increased power of our approach.

**Table 1.**
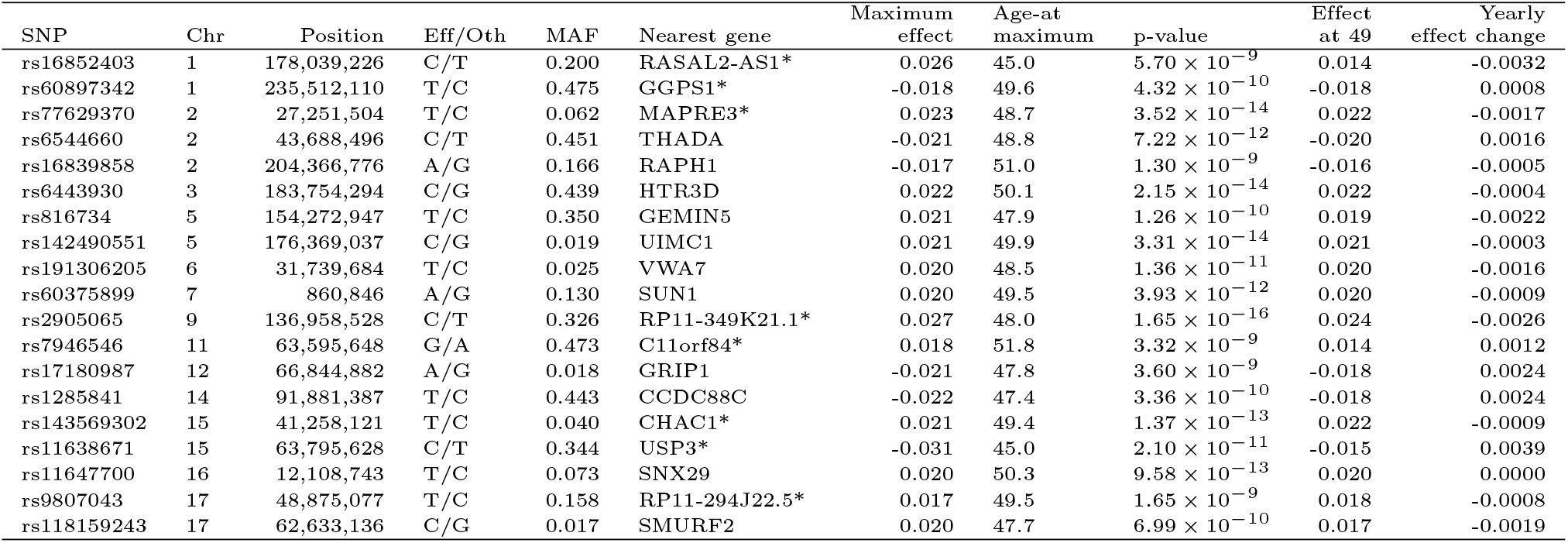
Previously undiscovered regions affecting age-at-menopause. For 8.7M SNPs, we determined the age at which there is the strongest evidence for an effect within the CAMP model. Then given the age identified for each SNP, we tested for significance at this age using the CAMP model results, and we obtained effect size and standard error estimates. The results were then LD clumped such that the index SNPs would have a p-value below 5 · 10^−8^ and SNPs could be added to a clump if they were 1Mb from the index SNP, they were correlated with *r*^2^ *>* 0.05 and they were nominally significant (*p <* 0.05). We then used the COJO method from the GCTA software (see Methods) to find clumps with independent signals by conducting a stepwise selection of index SNPs in a 1Mb window and we considered SNPs independent if they had a p-value below 5 · 10^−8^ in the joint model. To determine novelty, we then removed all the markers that had a correlation of *r*^2^ *>* 0.1 with a marker that had been previously found associated with age-at-menopause using the GWAS Catalog and LDtrait tool with the British in England and Scotland population. For the remaining SNPs, we conducted an additional literature review using the Phenoscanner database (see Methods) to find any previous associations with variants of interest or variants in LD. The remaining candidates for novel associations were then tested in the Estonian Biobank. Replication was defined as a p-value lower than 0.05 and the direction of the effect size same in both the original analysis and the replication analysis. The effect size estimates are reported on the log hazard scale. The column named, nearest gene, is mapped from the SNP using ANNOVAR software.

### Proportion of age-specific effects

For quantifying the existence of age-specific effects, we first test the null hypothesis of whether the slope term 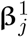 is different from 0, with a rejection of the null implying the existence of a time-varying genetic effect. Of the 245 UK Biobank associations, we find that 72% (176) show at least nominally significant (*p* < 0.05) age-specific effects within UK women (Figure 2c). Applying a more stringent criterion, 37 of these 176 associations, 15% of all associations, have a slope with a genome-wide significant p-value. In total, we find 63 regions that exhibit a genome-wide significant slope term in the UK Biobank (Tables 2, S1), and we replicate the age-specific effects for 20 regions in the Estonian Biobank (Table 2) yielding a replication rate of 32% (Figure 2c). These 20 variants have stronger effect sizes earlier in life that mostly decay toward zero after age 50 (Figure S5), making them early-ANM specific. Although 139 variants do not pass the threshold for genome-wide significance, they still indicate that for many regions previously identified as menopause-associated, the assumption of constant effect size (assumption of proportional hazards at the SNP) is generally invalid. Indeed, the 43 UK Biobank discovered variants with significant slope terms that did not replicate in the Estonian Biobank had effect size directions that were broadly concordant across studies (Fisher’s exact test *p* = 0.051, Table S1).

**Table 2.** Regions with a genome-wide significant age-specific effect on age-at-natural menopause replicated in the Estonian Biobank. For each SNP, we tested the significance of the slope parameter using the CAMP model. The results were then LD clumped such that the index SNPs would have a p-value below 5 · 10^−8^ and SNPs could be added to a clump if they were 1Mb from the index SNP, they were correlated with *r*^2^ *>* 0.05 and they were nominally significant (*p <* 0.05). We then used the COJO method from the GCTA software (see Methods) to find clumps with independent signals by conducting a stepwise selection of index SNPs in a 1Mb window and we considered SNPs independent if they had a p-value below 5 · 10^−8^ in the joint model. The candidates for significant slope were then replicated in the Estonian Biobank. Replication was defined as a p-value lower than 0.05 and the direction of the effect size same in both the original analysis and the replication analysis. The effect size estimates are reported on the log hazard scale. The column named, nearest gene, is mapped from the SNP using ANNOVAR software, a * in that column denotes intergenic regions; chromosome X nearest gene was determined by using the UCSC Genome Browser. The column named, strongest evidence p-value, indicates the p-value at the age when there is the strongest evidence for an effect.

| SNP | Chr | Position | Eff/Oth | MAF | Nearest gene | Yearly effect change | Slope p-value | Strongest evidence p-value |
| --- | --- | --- | --- | --- | --- | --- | --- | --- |
| rs6684319 | 1 | 39,334,988 | A/G | 0.310 | MYCBP | 0.0061 | $1.12 \times 10^{-19}$ | $9.63 \times 10^{-57}$ |
| rs185012833 | 2 | 62,779,457 | C/A | 0.003 | PSAT1P2* | -0.0033 | $6.03 \times 10^{-9}$ | $1.81 \times 10^{-6}$ |
| rs6760293 | 2 | 171,816,531 | A/T | 0.373 | GORASP2 | -0.0040 | $1.36 \times 10^{-9}$ | $7.33 \times 10^{-34}$ |
| rs12503643 | 4 | 185,746,088 | T/G | 0.399 | ACSL1 | 0.0049 | $3.05 \times 10^{-13}$ | $4.28 \times 10^{-61}$ |
| rs274722 | 5 | 6,718,668 | T/C | 0.406 | PAPD7 | -0.0040 | $1.06 \times 10^{-9}$ | $3.64 \times 10^{-25}$ |
| rs58400555 | 5 | 176,454,081 | T/A | 0.484 | ZNF346 | 0.0039 | $5.34 \times 10^{-9}$ | $3.21 \times 10^{-179}$ |
| rs2077491 | 6 | 31,606,376 | C/T | 0.473 | BAG6* | 0.0053 | $1.67 \times 10^{-15}$ | $1.06 \times 10^{-60}$ |
| rs728900 | 10 | 131,590,300 | A/T | 0.420 | RP11-109A6.3* | -0.0050 | $5.79 \times 10^{-14}$ | $4.19 \times 10^{-28}$ |
| rs75770066 | 12 | 66,704,225 | G/A | 0.033 | HELB | 0.0049 | $6.62 \times 10^{-12}$ | $1.58 \times 10^{-212}$ |
| rs11638671 | 15 | 63,795,628 | C/T | 0.344 | USP3* | 0.0039 | $3.62 \times 10^{-9}$ | $1.09 \times 10^{-10}$ |
| rs33650 | 16 | 11,978,769 | C/T | 0.385 | GSPT1 | 0.0042 | $3.28 \times 10^{-10}$ | $6.75 \times 10^{-65}$ |
| rs1433753 | 16 | 34,879,951 | T/C | 0.441 | RP11-14K3.1* | -0.0048 | $4.21 \times 10^{-13}$ | $2.26 \times 10^{-23}$ |
| rs8071278 | 17 | 41,193,910 | T/A | 0.335 | BRCA1* | 0.0048 | $5.01 \times 10^{-13}$ | $4.70 \times 10^{-50}$ |
| rs1991401 | 17 | 62,502,435 | G/A | 0.310 | DDX5 | -0.0048 | $2.51 \times 10^{-13}$ | $2.96 \times 10^{-36}$ |
| rs16960290 | 19 | 55,799,918 | T/C | 0.433 | BRSK1 | 0.0079 | $2.80 \times 10^{-32}$ | $1.11 \times 10^{-86}$ |
| rs117146677 | 19 | 55,833,868 | A/G | 0.009 | TMEM150B | -0.0053 | $1.50 \times 10^{-18}$ | $1.61 \times 10^{-43}$ |
| rs299163 | 19 | 56,321,414 | C/A | 0.066 | NLRP11 | 0.0042 | $4.26 \times 10^{-10}$ | $8.36 \times 10^{-17}$ |
| rs8124538 | 20 | 61,300,863 | A/G | 0.212 | SLCO4A1 | -0.0039 | $1.65 \times 10^{-9}$ | $3.63 \times 10^{-54}$ |
| rs6631137 | X | 30,665,762 | C/T | 0.316 | GK* | -0.0064 | $1.60 \times 10^{-22}$ | $5.43 \times 10^{-43}$ |
| rs67596711 | X | 152,638,744 | G/T | 0.499 | ZNF275* | 0.0055 | $1.28 \times 10^{-16}$ | $3.82 \times 10^{-26}$ |

**Figure 2.**
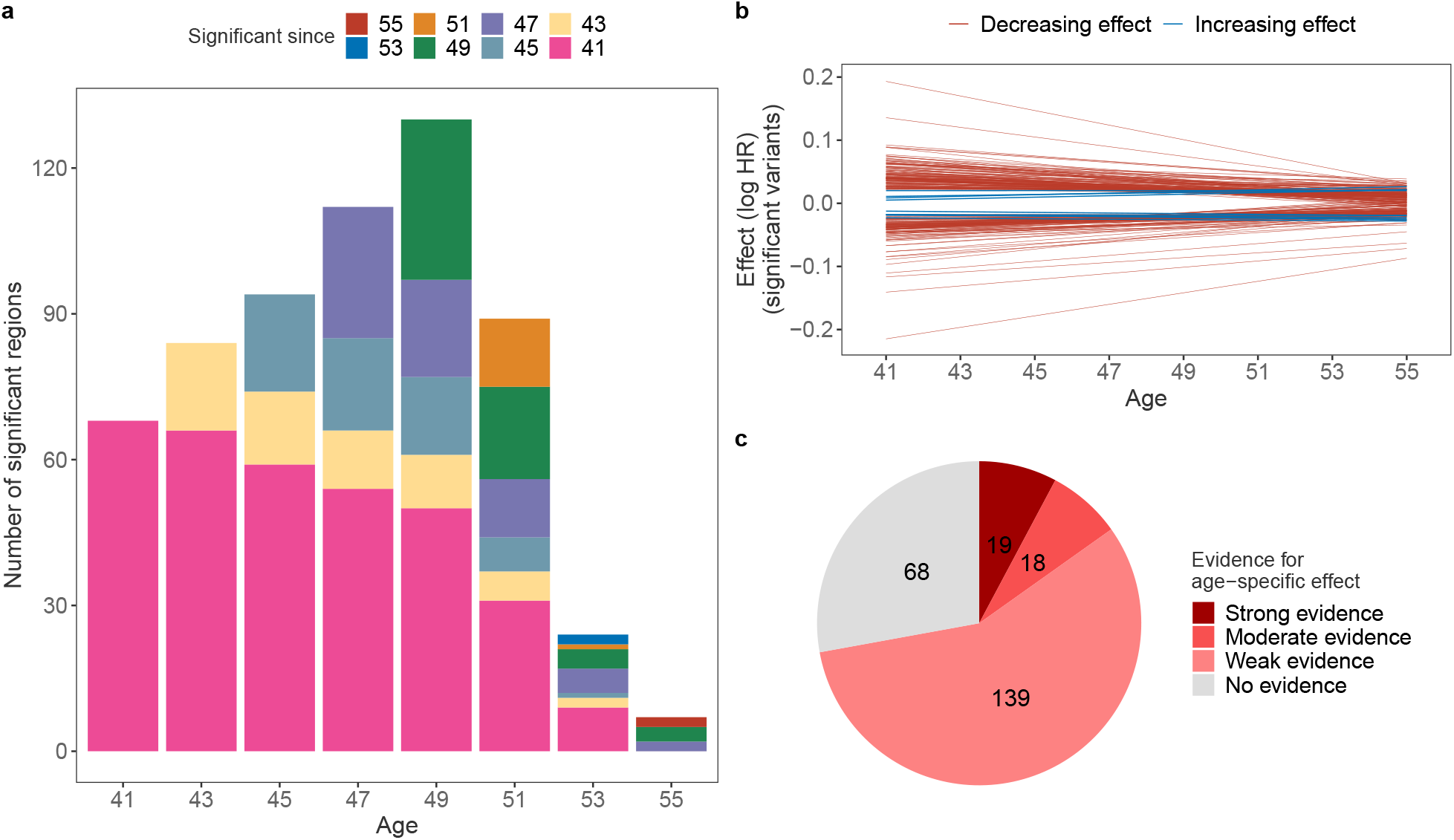
Age distribution of significant effects and effect size change. (a) We evaluated the significance of every SNP at each age on a grid from 41 to 55 and counted the number of significant hits replicated in the Estonian Biobank. The significant focal hits were mapped to consecutive ages, summarising the count since when the effects were significant; (b) Effect sizes (log HR) for 245 significant effects, the majority of variants have a larger absolute effect size at age 41 than at age 55, only nine variants have an increasing effect size. We observe that the model manages to capture the effect size change for many variants; (c) Classification of the menopause-associated variants by age-specific evidence by testing whether the slope parameter is equal to zero. Variants with weak evidence have a p-value lower than *p <* 0.05, moderate evidence requires *p <* 5 · 10^−8^, and strong evidence requires the slope to be significant also in the replication data set.

Second, as the model is age-specific, we test for significance at time points where the evidence is the highest within the intervals 41 to 55 (see Methods). For significant SNPs detected in the UK Biobank, the age distribution of maximum association evidence is concentrated between the ages of 43 and 51 (Figure S3). That is different compared to the maximum association evidence age distribution for all SNPs, which has thicker tails with nearly four times higher standard deviation even if the distributions have similar centres (median age of 51 and 49 for all SNPs and significant SNPs, respectively) (Figure S3). We observe that the number of regions affecting ANM changes considerably with the peak number of ANM-affecting regions observed at age 49 (Figure 2a). Moreover, we find that the period during which a particular region can significantly impact ANM varies considerably with only half of the significant associations at age 47 also significant at age 41. In general, we observe that effects tend to become insignificant with increasing age, with the drop in significance occurring at age 53, so that by age 55 only 8 loci have a genome-wide significant effect on age-at-menopause (Figure 2a). A similar result can be seen if we observe the distribution of ages when the evidence for the menopause effect is the strongest (Figure S3), as very few significant SNPs achieve the strongest association after age 51.

In contrast to most associations discovered at age 49, the general trend across 245 significant SNPs is that the effect size estimates shrink towards zero (Figure 2b). That might imply that the increase in the number of discoveries in the period 41 to 49 is instead due to the reduction in the standard error, and with a higher sample size, it could be possible to detect more associations already at earlier ages. Interestingly, only nine of the significant SNPs have a larger absolute effect size at age 55 than at 41. That is in line with many previous results reporting a reduction in relative genetic risks with the increase in age [10, 11]. Finally, we observe that there exists a stark difference between the effect size profiles of significant and insignificant effects (Figure S4) with a much narrower effect size distribution for the non-significant SNPs. Meanwhile, menopause-associated variants stand out as their effect size can change greatly across the period of interest.

Our analysis differs in one other key way from previous ANM genetic association studies. Here, we do not censor women who were placed on hormone replacement therapy (HRT). In survival models, declaring HRT individuals as censored makes the modelling assumption that age-at-HRT start and ANM are independent. They are clearly not as for women on HRT, there is a correlation between the age of HRT and ANM of 0.58 within the UK Biobank. For women who were placed on HRT prior to the recorded date of ANM, this correlation is stronger at 0.69. A Cox Proportional Hazards model for ANM including a categorical covariate of whether a woman was given HRT prior to menopause (1 if on HRT prior to menopause, 0 otherwise), shows that censoring for HRT prior to ANM, would significantly censor for earlier menopause (HR = 0.95, *p* = 6.08 · 10^−15^). Thus, censoring for HRT is not the optimal modelling choice and additionally, it results in the loss of 34,031 observations, reducing power. Nevertheless, we conduct a sensitivity analysis of our estimated effect sizes with and without adding HRT as a time-varying covariate to the CAMP model, at the 312 top loci identified within our study (Figure S6). We find very strong concordance of effect sizes across loci (Figure S6), highlighting that in practice these different modelling choices have no detectable impact on the leading SNP association findings.

### Properties of novel and age-specific genetic associations

We conduct a number of follow-up analyses to support our age-specific association results. First, we test for significant enrichment of the summary statistics generated by our approach for each age group. For all categories showing significant enrichment after Bonferroni multiple testing correction, we find that their significance does not hold across all age groups (Figure 3 and Table S3 to Table S10).

**Figure 3.**
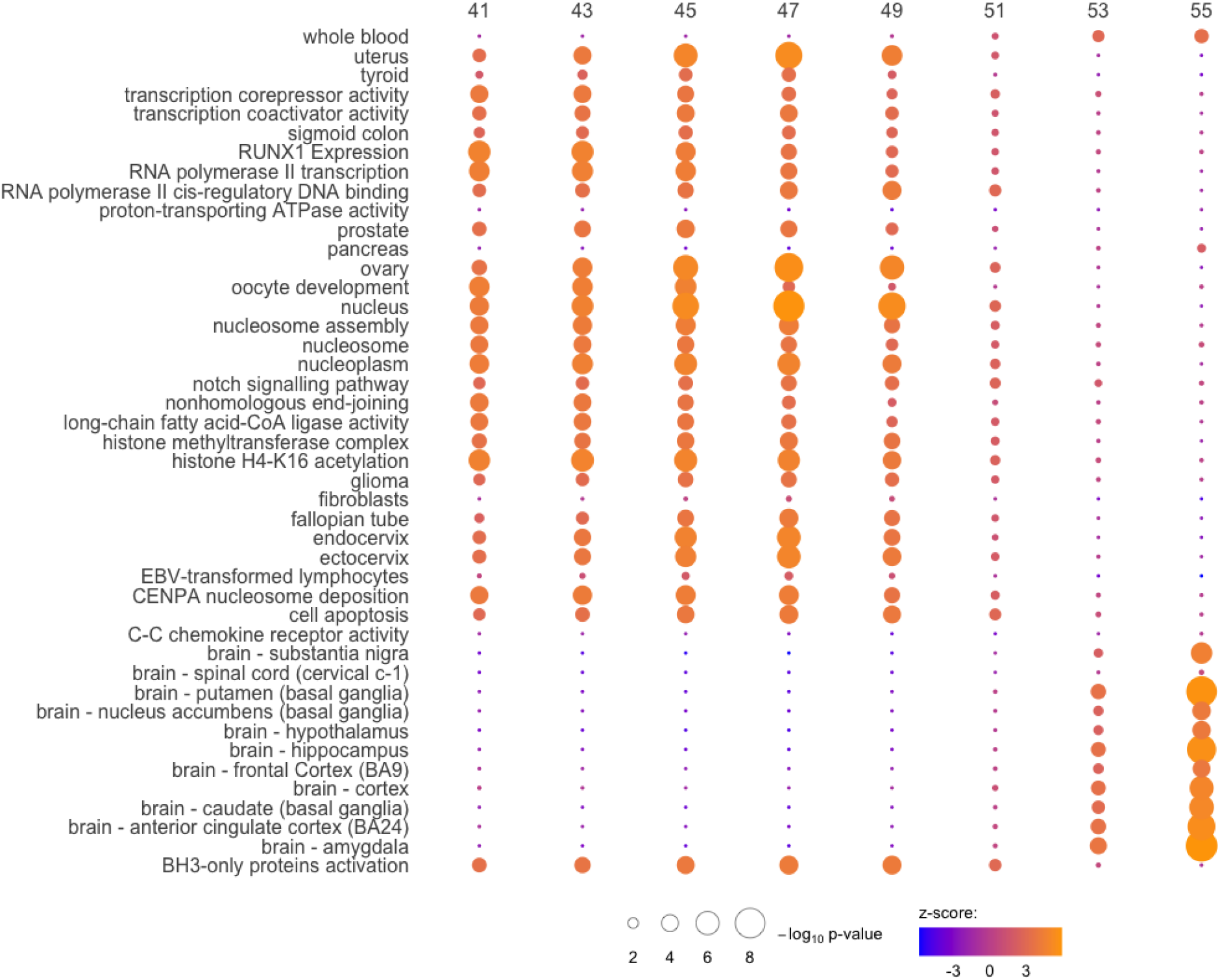
Age-specific enrichment of genetic associations across multiple genomics resources. We evaluated the significance of every SNP at each age on a grid from 41 to 55 and from the resulting summary statistics we tested for enrichment across multiple genomics resources. Circle circumference gives the −log_10_ p-value and the colour gives the enrichment z-score calculated from the Downstreamer software. GTEx tissue-specific expression and GO terms are given on the y-axis for annotations with genome-wide significance after multiple testing correction at one or more age groups. Full results are given in Table S3 to Table S10.

Effect sizes for ANM between the ages of 41 to 49 were enriched in genes differentially expressed in the uterus, thyroid, prostate, ovary, fallopian tubes, and cervix within the GTEx consortium data (Figure 3). Additionally, we find enrichment between the ages of 41 and 49 for KEGG pathway NOTCH signalling associated with cell proliferation and death, the GO terms for an intrinsic pathway for apoptosis, and BH3-only proteins (Figure 3). In contrast, associations with variation in ANM for individuals older than 51 were all enriched for genes with differential expression in several brain regions within the GTEx data, with no evidence for enrichment in reproductive tissues (Figure 3). These results suggest that genetic effects may differ across the age range.

Our next follow-up analysis used LD Score regression, where we find that genetic correlations across ages are significantly less than 1 (Figure 4a). Genetic correlations of ANM and other phenotypes were also largely age-dependent (Figure 4b). Note here that effect size estimates for ANM are calculated on the menopause hazard scale and thus a positive correlation estimated by LD Score regression would refer to a high hazard of ANM (earlier ANM) corresponding to high trait values, in other words, the observed value of ANM and the trait are in fact negatively correlated. Thus, to ease interpretation we flip the sign of the estimated correlation to display the genetic correlation of the observed values of ANM and each trait.

**Figure 4.**
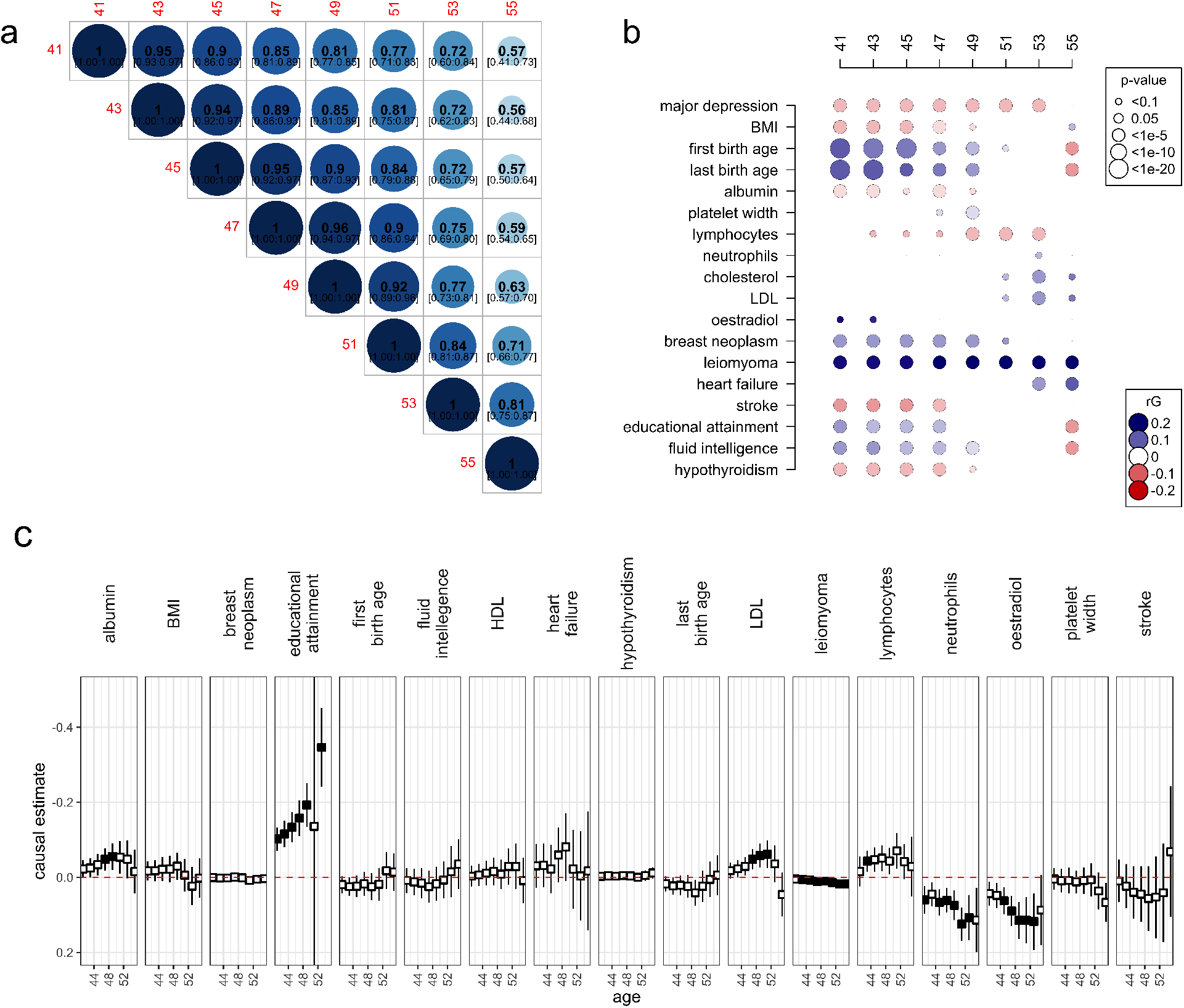
Age-specific genetic correlations and causality of ANM and health-related outcomes. We evaluated the significance of every SNP at each age on a grid from 41 to 55 and from the resulting summary statistics we tested for genetic correlations among (a) age groups and (b) among observed ANM values at the age groups and 100 other health-related indicators and outcomes, using LD Score regression. In (b), we present correlations for outcomes with evidence of a significant non-zero genetic correlation at one age group or more. In (c), we present results from inverse-weighted Mendelian Randomization that estimates the potential causal relationship between ANM and outcomes where a significant genetic correlation was found in (b) across ages. Black boxes depict ages for which significant causal estimates were found. Full results from a range of Mendelian Randomisation models are given in Table S11.

Between 41 and 49 years, we find a significant positive genetic correlation of observed ANM values with age at last birth and age at first birth (Figure 4b), implying a genetic relationship between later reproduction and later ANM, for women whose ANM is earlier than average.

Of significant note, genetic propensity for breast cancer was significantly associated with a later ANM (Figure 4b), before age 51. This supports previous evidence, where genetically mediated delays in ANM were found to increase the relative risks of several hormone-sensitive cancers [8]. Additionally, evidence linking exposure to high levels of estrogen hormones to an increased risk of breast cancer is supported by a significant positive genetic correlation of ANM and oestradiol levels for women of early menopause, implying a genetic propensity for high estrogen levels associated with a genetic propensity for a later ANM (Figure 4b). Furthermore, we find that a high genetic risk for leiomyoma is consistently associated with later ANM (Figure 4b). Together, with our enrichment results presented above (Figure 3) showing early ANM genetic associations are enriched for genes differentially expressed female reproductive organ, oocytes, and DNA damage repair mechanisms, our findings suggest that at the genetic level, breast cancer risk, hormone levels and ANM are intrinsically linked prior to age 50.

We also find significant genetic correlations implying that genetically mediated later ANM is correlated with a lower genetic predisposition to hypothyroidism, stroke, major depression, blood albumin levels, and obesity prior to age 51 (Figure 3). For women at later ages, we find positive genetic correlations of ANM with cholesterol, LDL, obesity, and heart failure, implying that later ANM is correlated with an increased genetic predisposition for these metabolic-associated health measures (Figure 3).

We find a significant positive genetic correlation of ANM values with both educational attainment and fluid intelligence between 41 and 49 years (Figure 4b), implying a genetic relationship between later reproduction, higher education and later ANM, for women before the age of 50. Interestingly, these genetic correlations also significantly change in the sign for women whose ANM occurred after age 53, with a significant negative genetic correlation of ANM with educational attainment and fluid intelligence (Figure 4b), implying delayed reproduction and high educational attainment are associated with reproductive senescence post-age 50.

In a further follow-up analysis, we used Mendelian Randomisation (MR), which utilizes the randomized inheritance of genetic variations in the population to estimate the potential causal effect a modifiable risk factor or exposure has on a health-related outcome of interest. We used menopause at different ages as an exposure in five different MR methods (Weighted median, Inverse variance weighted, Simple mode, Weighted mode, and MR-Egger, see Table S11) found in the ‘TwoSampleMR’ R package. Note here again that effect size estimates for ANM are calculated on the menopause hazard scale and thus to ease interpretation, we flip the sign of the estimated potential causal effect to give values on the observed ANM scale. When repeating the analysis for each varying age of our exposure, we find changes in the magnitude of the potential causal effect with age for educational attainment, leiomyoma, oestradiol, and neutrophil count (Figure 4c).

Finally, we highlight notable examples of the novel replicated associations with significant slope terms, such as chr11:63,595,648 which is downstream of the SPINDOC gene, where menopause genetic risk increases with age, with the highest effect size at later age groups. Also, chr15:63,795,628 which is upstream of the USP3 gene where the menopause association disappears with increasing age. Both of these associations were previously suggestively associated with age-at-menopause, but passed the significance threshold in the UK Biobank and replicate in the Estonian Biobank using our proposed model.

## Discussion

Taken together, we find that the majority of ANM genetic associations display some form of age-specificity in their effects. In turn, that translates into the associations being differentially enriched in different biological pathways across ages, which then leads to different genetic associations of ANM and other health indicators and outcomes depending upon the timing of ANM, with different potential statistical causal relationships.

We find evidence that prolonged and delayed reproduction are genetically associated with reproductive senescence post-age 50 as the genetic correlations significantly change in the sign for women whose ANM occurred after age 53, with a significant negative genetic correlation of ANM with age at last birth and age at first birth (Figure 4b). Similarly, we find genetic correlations between ANM and educational attainment or fluid intelligence significantly turn negative for women whose ANM occurred after age 53. The latter patterns of changing genetic correlation may simply reflect reproductive choices of the timing and number of children made in the human population that are associated with levels of educational attainment.

Complementing the results from genetic correlations, our enrichment analysis results (Figure 3) show early ANM genetic associations being enriched for genes differentially expressed in female reproductive organs, oocytes, and DNA damage repair mechanisms. Hence, our findings suggest that at the genetic level, breast cancer risk, hormone levels and ANM are intrinsically linked prior to age 50. The patterns observed in the MR analyses largely reflect those of the genetic correlations described above, but here we find little evidence for a causal relationship between ANM and breast cancer, nor heart failure, age at first or last birth, or hypothyroidism (Figure 4c). That implies that genetic correlation estimates likely reflect reverse causation or the presence of heritable confounders of the trait pairs.

Our enrichment analysis findings support a link between DNA damage repair genes and repair and surveillance for the development of oocytes for early-ANM women. The size of the initial oocyte pool at birth, along with the rate of atresia, influences the age at which the oocyte pool is depleted. The meiosis that occurs in oocytes necessitates programmed double-stranded breaks (DSBs) that must be repaired through the homologous recombination pathway, with oocytes that do not properly repair DSBs after this first phase of meiosis undergoing apoptosis. Here, early-ANM-associated common variants are enriched at loci harbouring genes involved in the DNA repair, and replication checkpoint processes, such as RNA polymerase II, histone methyltransferase complex and histone acetylation (Figure 3). One-carbon metabolism has the ability to regulate the estrus cycle and modulate the initiation of reproductive senescence through the loss of methyl-donor production needed to properly maintain the epigenome. Our results support the existence of this mechanism as early-menopause associations are enriched in pathways associated with the hypothalamic-pituitary-gonadal (HPG) axis and with methylation in the nucleosome, with later-menopausal genetic associations showing no evidence of enrichment in these pathways (Figure 3). In humans, it has been suggested that postmenopausal women exhibit accelerated ageing compared with premenopausal women of the same biological age [12]. However, the cause-effect relationship between epigenetic changes and reproductive senescence remains unclear and our results imply early-ANM women may have a methylation pattern associated with one-carbon metabolism that differs from the general population. Generally, our follow-up analyses support previous studies [9, 13], but we demonstrate that almost all underlying pathways associated with variation in ANM act in an age-specific manner.

There are several important caveats to our study. Firstly, we have assumed that the effect size can only change linearly with age, whereas in reality, they could consist of more complicated patterns that could be captured with piece-wise exponential models. However, introducing many more parameters on a genome-wide scale would lead to a high multiple-testing burden, potentially hampering the capability to detect the actual signal. Furthermore, especially for traits with a moderate range of values (90% of the observed ANM happen between ages 45 to 55, Figure S1), introducing many parameters could lead to overfitting. Therefore, we find that, especially in the context of traits such as age-at-menopause, assuming a linear effect change is a suitable compromise between the added value of learning new information about effect change and limiting the model complexity without damaging the statistical power. Nevertheless, the analyses presented here represent a first step, and we encourage specifying different functional forms for the effect size, preferably for traits with a broader range of values or a reasonable prior guess.

Second, the current implementation of the model is not computationally efficient and to handle the computational burden we have utilised the computational resources of two universities to produce these results. Our objective was simply to simply highlight the existence of changing genetic relationships between phenotypes and health outcomes across the lifespan. Although it is possible to make marginal analyses embarrassingly parallel, it is inherently time-consuming to fit an age-specific Cox proportional hazards model. Scaling the inference requires new research into novel algorithms for computationally heavy high-dimensional statistical problems of this kind.

Finally, our study focused only on European ancestry individuals in the UK and Estonian Biobanks, and future analyses must take into account populations with more diverse ancestries to get a fuller picture of the genetic architecture of age-at-menopause across the globe. This requires research into statistical models that are capable of learning both shared and unique age-dependent effect sizes across populations and it requires large-scale data to be collected from worldwide populations.

In summary, we propose a novel analysis approach for GWAS of age-at-onset phenotypes, using a two-stage MLMA model, where marker effect sizes are estimated using an age-specific Cox proportional hazards model. Our approach provides a better understanding of the genetic basis of age-at-menopause and applies to any form of time-to-event phenotype.

## Methods

### Marginal Age-specific Mixed Cox Proportional hazards model

Following the success of many GWASs, more attention has been attributed to better characterising the SNP effect behaviour under different environmental conditions, leading to genotype-covariate analyses [14]. For a continuous trait, one of the simplest ways to model this type of interaction would be to include a linear interaction term. For example, to estimate the impact of age on SNP effects, we could write the model as:

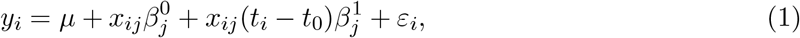

where (for individual *i* and SNP *j*) *y*_*i*_ is a continuous trait, *µ* is the intercept, *x*_*ij*_ is the SNP value, 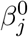 is the SNP effect at time *t*_0_, *t*_*i*_ is the age when *y*_*ij*_ is measured, 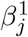 is the linear effect of age and *ε*_*i*_ is the residual variance. However, for age-at-onset phenotypes, the model as specified in Equation 1 would not be identifiable since *y*_*i*_ = *t*_*i*_. Previous studies have proposed to analyse age-specific effects by splitting time scales into non-overlapping intervals. Individuals who have the event in a future interval are treated as right censored, with individuals who have had an event in a previous interval excluded from the analysis. For example, this idea has been suggested by Joshi et al. [15], where time intervals of 40-75 and 75+ were used. Although it is correct to conduct the analysis in such a way, it requires defining intervals that could be seen as an arbitrary choice, with ill-defined intervals leading to an incomplete understanding of the effect size distribution. Furthermore, this type of modelling will not scale well with the added number of intervals as each interval requires an additional parameter. Therefore, we propose a Cox PH model that allows specifying a functional shape for age-specific effects. To estimate the marginal effect of SNP *j* in chromosome *k*, the general form for Cox proportional hazards model for each SNP *j* ∈ 1, …, *M* is

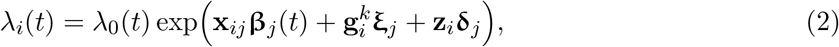

where λ_0_(*t*) is the baseline hazard, *i* denotes the *i*th individual, **x**_*ij*_ is the standardised *j*th marker value, 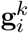 is the genetic predictor from all other chromosomes other than the SNP *j* is located at, **ξ**_*j*_ is its corresponding effect when estimating marker *j*, **z**_*i*_ is the summarised covariate value and **δ**_*j*_ is its corresponding effect when estimating marker *j*. **β**_*j*_(*t*) is the effect size change function for SNP *j* and we define it in two ways. The effect size function **β**_*j*_(*t*) should be defined such that its domain is the set of positive real numbers and it is (piecewise) differentiable. Here, we define the effect size function as a linear function in age

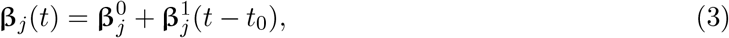

where 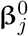 is the intercept term, effectively estimating the effect size at time *t*_0_ and 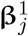 is the slope showing how much the effect changes each year. It is possible to define any other parametric shape, for example, the exponential decay function could be a natural choice. However, in the first experiments on real data, the linear effect size change gave a higher likelihood compared to exponential decay models. Hence, we decided to resort to the linear effect size model that is easier to interpret and the variance function can be represented without Taylor expansion-based approximations. Given the effect size function definition, we can calculate the variance at each time point *t* as

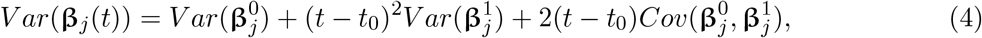

where 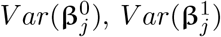 and 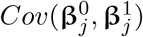 can be estimated from the Hessian of the Cox model. We use the linear effect change definition from equation 3 to test whether there exists a change in the effect size across the lifespan. As including the genetic values from other chromosomes shares properties with mixed modelling, we are going to refer to this model as the CAMP model (Cox Age-specific Mixed Proportional hazard).

At each time point *t* we can define the test statistic function 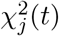 for SNP *j* as the square of the ratio of effect size and the standard error of the effect size

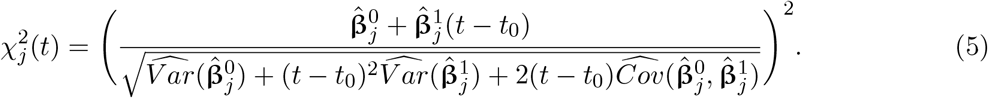

All that remains is to estimate 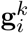, the genetic predictor from all other chromosomes other than the SNP *j*. To do this, we use a BayesW model [1] which assumes that for an individual *i* the age-at-onset of a disease *y*_*i*_ has Weibull distribution, with a re-parameterisation of the model to represent the mean and the variance of the logarithm of the phenotype as

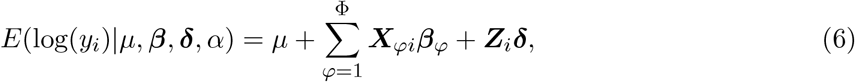

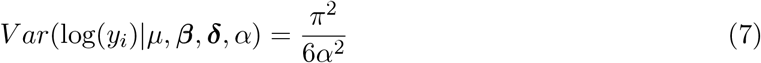

where ***X***_*φ*_ is a standardised genotype matrix containing SNPs allocated to group *φ, µ* is an intercept, ***β***_*φ*_ is the vector of SNP effects in group *φ*, ***Z***_*i*_ are additional covariates (such as sex or genetic principal components), ***δ*** are the additional covariate effect estimates and *α* is the Weibull shape parameter. For each group, we assume that ***β***_*φ*_ are distributed according to a mixture of Gaussian components with mixture-specific proportions ***π***_*φ*_ and mixture variances 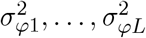 and a Dirac delta at zero which induces sparsity:

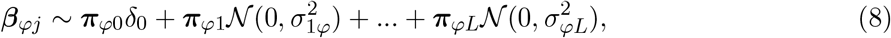

where *L* is the number of mixture components. We estimated the hyperparameters such as genetic variance and prior inclusion probability by grouping markers into MAF-LD bins as recent theory suggests this yields improved estimation [16] and here, 20 MAF-LD groups that were defined as MAF quintiles and then quartiles within each of those MAF, quintiles split by the LD score. The cut-off points for creating the MAF quintiles were 0.006, 0.013, 0.039, 0.172; the cut-off points for creating LD score quartiles were 2.11, 3.08, 4.51 for the first; 3.20, 4.71, 6.84 for the second; 4.70, 6.89, 9.94 for the third; 7.65, 11.01, 15.70 for the fourth and 10.75, 15.10, 21.14 for the fifth MAF quintile, exactly as in the age-at-menopause analysis by Ojavee et al. [1]. The posterior mean BayesW model estimates of ***β***_*φ*_ are then used to create 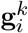, the genetic predictor from all other chromosomes other than the SNP *j*. This gives a two-step LOCO (leave-one-chromosome-out) approach, where first a BayesW model is used to estimate the genetic predictor and then a marginal age-specific Cox proportional hazards model is used for the second step. Next, we discuss how we can conduct significance testing in the second step in an efficient manner, whilst ensuring that type I error is bounded below the fixed threshold *α* even with a more complex model.

### Significance testing

We demonstrate that to limit type I error rate below *α* and given the null hypothesis of no effect at SNP *j* 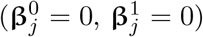, it is sufficient to compare the test statistic 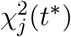 with the 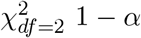 quantiles at any time point *t*\*.

We will naturally assume that 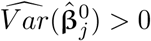 and 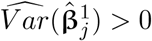. As 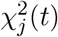 is twice differentiable, it is possible to find its respective first and second derivative. This will give us two extreme points *t*\* at which the 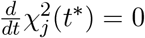. Then, the *χ*^2^-score has a local maximum if 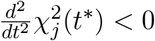 or the *χ*^2^-score has a local minimum weakest if 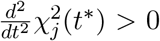. It can be shown that the function 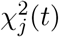 has two extreme points located at

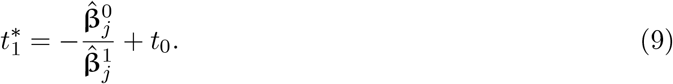

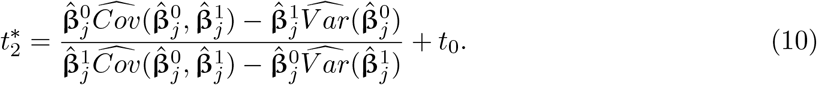

Using the second derivative, it can be shown that 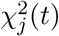 will always have a (global) maximum at 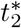 and a (global) minimum 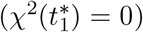 at 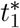. We find that in the limiting cases test statistic is testing the significance of the slope 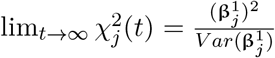 and 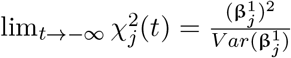. As the domain of 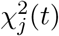 is the set of positive real numbers and there are no breakpoints in the function, then 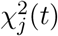 is bounded within interval 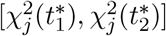.

We are especially interested in the distribution of the maximum possible *χ*^2^-statistic 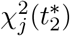 under the null hypothesis that both 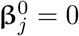 and 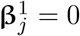.

#### Lemma 1

*Under the null hypothesis that both* 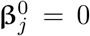 *and* 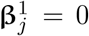, *the chi-squared statistic evaluated at the maximum point* 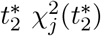 *follows a* 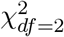 *distribution:*

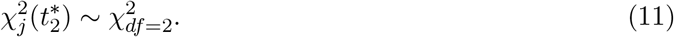

*Proof*. We define 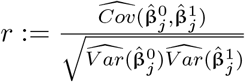 and we express 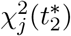 such that it would be a sum of two uncorrelated random variables.

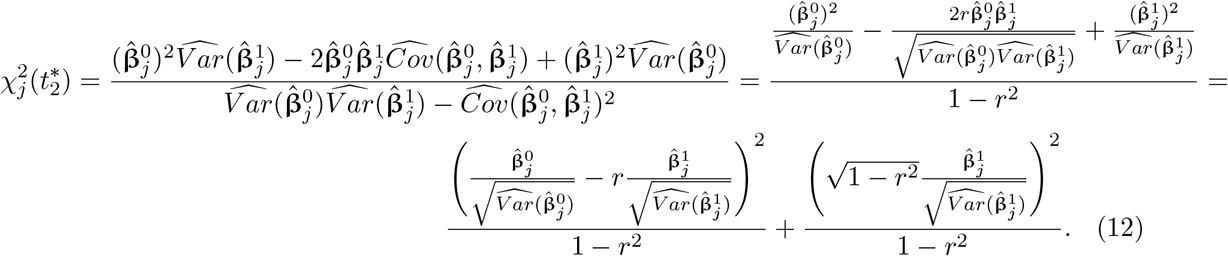

Under the null hypothesis of 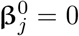 and 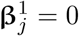 we know that 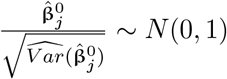 and 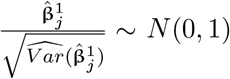 and therefore

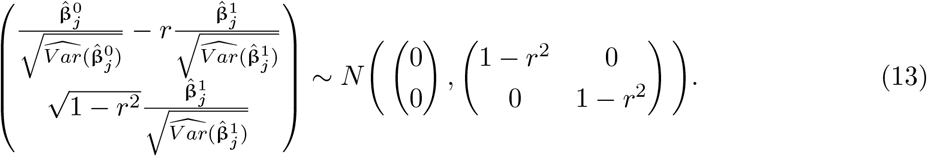

The last result implies that under the null of 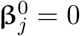 and 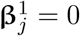 the equation 12 is a sum of two uncorrelated standard Gaussian random variables squared which means that 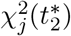 is from the chi-squared distribution with degrees of freedom of 2.

This naturally gives us a rule for hypothesis testing at time 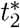. If the test fails to disprove the null hypothesis at time 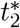, it will fail to disprove the null hypothesis at any possible *t*. If the test accepts the alternative hypothesis, it means that there must exist an interval (or at least one point) at which the variable has an effect on the phenotype.

Furthermore, we can show that the quantiles of 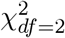 distribution result in a stringent enough test at any time point.

#### Lemma 2

*Suppose that we have estimated effect sizes* 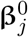 *and* 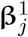 *from a linear effect change model* 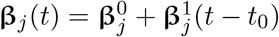 *and that the null hypothesis of* 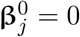 *and* 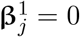 *holds. Then, for every time point t, the probability of type I error (α) is bounded when using the* 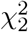 *distribution* 1 − *α quantile as a critical value*.

*Proof*. To prove the lemma, we need to demonstrate that 1 − *α* quantile of 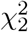 distribution (*q*_1−*α*_) is greater than 1 − *α* quantile of *χ*^2^(*t*) at any time point *t* under the null hypothesis 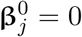 and 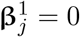. Suppose that the maximum test statistic value is achieved at 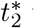 with value 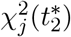.

We suppose in contradiction that under the null hypothesis there exists some time point 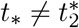 at which the distribution of 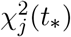 would have a higher 1 − *α* quantile value 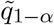 than the 1 − *α* quantile of 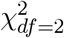 distribution *q*_1−*α*_:

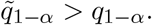

Given this, we can write the following inequalities

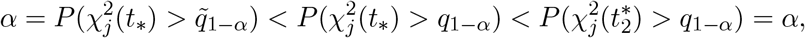

where the first inequality follows from the contradiction and the second inequality from the fact that 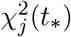 is the maximum possible value of the test statistic. The inequalities result in a contradiction which therefore proves the lemma.

An important corollary of this result is that we can use 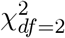 quantiles to do statistical testing at any time point and doing tests at (many) different time points will not increase type I error. For example, we can simultaneously test the significance of the slope (corresponds to 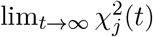) and significance at 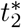 (using 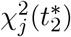 at the 1 − *α*-quantile of 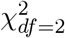 distribution while limiting the type I error at *α* as given the effect and variance estimates.

### UK Biobank data

We first restricted our analysis to a sample of European-ancestry UK Biobank individuals. To infer ancestry, we used both self-reported ethnic background (UK Biobank field 21000-0), selecting coding 1, and genetic ethnicity (UK Biobank field 22006-0), selecting coding 1. We projected the 488,377 genotyped participants onto the first two genotypic principal components (PC) calculated from 2,504 individuals of the 1,000 Genomes project. Using the obtained PC loadings, we then assigned each participant to the closest 1,000 Genomes project population, selecting individuals with PC1 projection < absolute value 4 and PC2 projection < absolute value 3. Samples were also excluded based on UK Biobank quality control procedures with individuals removed of (i) extreme heterozygosity and missing genotype outliers; (ii) a genetically inferred gender that did not match the self-reported gender; (iii) putative sex chromosome aneuploidy; (iv) exclusion from kinship inference; (v) withdrawn consent. We used genotype probabilities from version 3 of the imputed autosomal genotype data provided by the UK Biobank to hard-call the genotypes for variants with an imputation quality score above 0.3. The hard-call-threshold was 0.1, setting the genotypes with probability ≤ 0.9 as missing. From the good quality markers (with missingness less than 5% and p-value for Hardy-Weinberg test larger than 10^−6^, as determined in the set of unrelated Europeans), we selected those with minor allele frequency (MAF) > 0.0002 and rs identifier, in the set of European-ancestry participants. We then took the overlap with the Estonian Biobank data described below to give a final set of 8.7 million SNPs using both autosomal chromosomes and the X chromosome. This provides a set of high-quality SNP markers present across both discovery and prediction data sets.

We created the phenotypic data of age-at-menopause similarly to [1]. We used UKB field 3581 to obtain the time if available, and we excluded from the analysis 1) women who had reported having and later not having had menopause or vice versa, 2) women who said they had menopause but there is no record of the time of menopause (UKB field 2724), 3) women who have had a hysterectomy or the information about this is missing (UKB field 3591), 4) women whose menopause is before age 33 or after 65. Within the UK Biobank data, there were a total of 173,424 unrelated (only one person kept from second-degree or closer relative pairs) European ancestry women, out of which 125,697 had experienced menopause and 47,727 had not had menopause based on data field 2724. For computational convenience when conducting the joint BayesW analysis we created an additional subset of markers by removing markers in very high LD, through the selection of the highest MAF marker from any set of markers with LD *R*^2^ ≥ 0.8 within a 1Mb window. These filters resulted in a data set with 173,424 individuals and 2,174,071 markers for the first-step estimation of the LOCO genetic predictors and then in the second-step age-specific Cox Proportional hazards we analysed 8.7 million SNPs using both autosomal chromosomes and the X chromosome.

### Estonian Biobank data

To replicate the findings we used the Estonian Biobank with 70,082 women (22,740 with menopause, 47,342 without menopause). In the Estonian Biobank data, there were 195,432 individuals genotyped on Illumina Global Screening (GSA) arrays, which were imputed to an Estonian reference created from the whole genome sequence data of 2,244 participants [17]. From 11,130,313 markers with imputation quality score > 0.3, we selected SNPs that overlapped with those selected in the UK Biobank as described above using the same SNP sets for the first and second steps of the analyses.

### Analysis of age-at-menopause

We investigated the three questions as proposed in Figure 1: 1) we checked the interval at which there is a significant effect, 2) we tested the significance at ages at which the variants had the most evidence for an effect, 3) we tested for the existence of an age-specific effect.

### Interval at which variant is significant

We evaluated the test statistic function using Equation 5 at ages 41, 43, 45, 47, 49, 51, 53, 55 and calculated the p-values using the 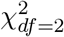 distribution quantiles as suggested by our theory above. On these results, at each age, we applied the clumping procedure (using plink 1.9 [18]) with a window size of 1Mb, LD threshold of *r*^2^ = 0.05, p-value threshold for index SNPs of *p* = 5 · 10^−8^ and no p-value threshold for other SNPs belonging to a clump of an index SNP. To detect clumps with an independent signal, we applied the COJO procedure [19] implemented in GCTA software [20] with a window size of 1Mb and the SNPs were considered independent if the p-value in the joint model was less than 5 · 10^−8^. The independent index SNPs from the COJO analysis were then replicated at each age in the Estonian Biobank data, with replication defined as a p-value lower than 0.05 and the same effect size estimate sign as in the discovery analysis. To check the period during which a SNP has a significant effect (Figure 2a), we checked whether the same SNP or a SNP in the same clump also has an effect in the consecutive grid point. Specifically, we took all the significant independent and replicated SNPs at ages 41 and 43, and then we checked if the index SNPs of age 41 mapped directly to an index SNP at age 43 or a clump of an index SNP. Then, we compared the ages 43 and 45, and iteratively so forth until 55.

### Testing at age with the maximum effect evidence

We tested the significance at ages at which the variants had the most evidence for an effect to understand the total number of significant effects and possibly identify novel loci. Furthermore, we defined a period of interest between ages 45 and 52, so the significance would only be evaluated during this period. That was done to avoid over-interpretation of the linear effect at uncommon high or low ages. Therefore, using Equation 10 we first calculated the age 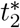 at which the test statistic achieves the highest value, and secondly, we evaluated the chi-squared function (Equation 5) at that age 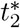. If there time point 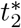 was outside of the interval [45,52], then we instead evaluated the function at the time point where the function 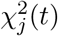 achieved its maximum within the interval (either age 45 or 52). We compared the chi-squared statistics with the 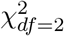 distribution to calculate the p-values. We applied a similar procedure to the previous case. First, we applied the clumping procedure (using plink 1.9 [18]) with a window size of 1Mb, LD threshold of *r*^2^ = 0.05, p-value threshold for index SNPs of *p <* 5 · 10^−8^ and no p-value threshold for other SNPs belonging to a clump of an index SNP. Secondly, to detect clumps with an independent signal, we applied the COJO procedure [19] implemented in GCTA software [20] with a window size of 1Mb and the SNPs were considered independent if the p-value in the joint model was less than 5 · 10^−8^. We then checked the independent and significant SNPs from the COJO analysis for previous association signals. We removed all the markers that had a correlation of *r*^2^ *>* 0.1 with a marker that had been previously found associated with age-at-menopause using the GWAS Catalog (published until April 2022) and LDtrait tool with the British in England and Scotland population. Furthermore, we specifically compared our candidate set for the significant SNPs reported by Ruth et al. [9], removing the markers or the markers with a correlation *r*^2^ *>* 0.1 reported by them. Finally, we checked our candidate set with the Phenoscanner database [21, 22] to find any previous associations with variants of interest or variants in LD. The index SNPs not removed by the three filters were then replicated at age with maximum evidence in the period [40,55] in the Estonian Biobank data, with replication defined as a p-value lower than 0.05 and the same effect estimate sign as in the discovery analysis.

### Testing for an age-specific effect

To verify whether there is a genome-wide significant age-specific effect for every SNP *j*, we checked if the slope parameter is significantly different from 0

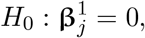

where 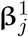 is estimated from the model specified in Equation 2. The chi-squared statistic was calculated as the squared ratio of the slope size estimate and standard error estimate. As this quantity naturally corresponds to 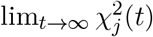 where 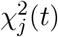 is defined as in Equation 5, we compare the test statistic again with the 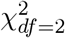 distribution quantiles to get the p-values. Similarly to two previous cases, we first applied the clumping procedure (using plink 1.9 [18]) with a window size of 1Mb, LD threshold of *r*^2^ = 0.05, p-value threshold for index SNPs of *p <* 5 · 10^−8^ and no p-value threshold for other SNPs belonging to a clump of an index SNP. Secondly, to detect clumps with an independent signal, we applied the COJO procedure [19] implemented in GCTA software [20] with a window size of 1Mb and the SNPs were considered independent if the p-value in the joint model was less than 5 · 10^−8^. The independent index SNPs from the COJO analysis were then replicated in the Estonian Biobank data, with replication defined as a p-value lower than 0.05 and the sign of the slope the same as in the discovery analysis. We classified the SNPs with different levels of evidence of age-specific effects. SNPs with a p-value below the nominal significance threshold (*p* < 0.05) are said to have at least weak evidence for an age-specific effect; variants with a p-value below the genome-wide significance threshold (*p <* 5 · 10^−8^) are said to have at least moderate evidence for an age-specific effect; variants with moderate evidence that also replicate in the Estonian Biobank are considered to have strong evidence for an age-specific effect. The variants that do not fall under these three categories are said to have no evidence for age-specific effects.

### Enrichment analysis

We used recently presented Downstreamer software to identify genes connected to our association study results through gene expression and to identify enriched pathways. We calculated test statistics using Equation 5 at ages 41, 43, 45, 47, 49, 51, 53, 55 and calculated the p-values using the 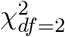 distribution quantiles as suggested by our theory above.

Downstreamer implements a strategy that accounts for LD structure and chromosomal organization, operating in two steps. In the first step, gene-level prioritization scores were calculated for each age group’s summary statistics and a null distribution. This aggregates p-values per variant into a p-value per gene while accounting for local LD structure. We aggregated all variants within a 25-kb window around the start and end of a gene using the non-Finnish European samples of the 1000 Genomes (1000G) project, Phase 3 to calculate LD. We calculated these GWAS gene pvalues for all 20,327 protein-coding genes (Ensembl release v75). The gene p-values were then converted to z-scores for use in subsequent analysis. These are referred to as GWAS gene z-scores. To account for the long-range effects of haplotype structure, which results in genes getting similar gene z-scores, we use a generalized least-squares (GLS) regression model for all regressions done in Downstreamer. The GLS model takes a correlation matrix that models this gene–gene correlation. To calculate this correlation matrix we first simulated 10,000 random phenotypes by drawing phenotypes from a normal distribution and then associating them to the genotypes of the 1000G Phase 3 non-Finnish European samples. We used only overlapping variants between the real traits and the permuted GWASs to avoid biases introduced by genotyping platforms or imputation. We then calculated the GWAS gene z-scores for each of the 10,000 simulated GWAS signals as described above. Next, we calculated the Pearson correlations between the GWAS gene z-scores. As simulated GWAS signals are random and independent of each other, any remaining correlation between GWAS gene z-scores reflects the underlying LD patterns and chromosomal organization of genes. An additional 10,000 GWASs were simulated to empirically determine enrichment p-values and, finally, we used an additional 100 simulations to estimate the FDR of Downstreamer associations.

In the second step, the gene-level prioritization scores were associated with the co-regulation matrix and pathway annotations. We used a previously generated co-regulation matrix that is based on a large multi-tissue gene network. In short, publicly available RNA-seq samples were downloaded from the European Nucleotide Archive (https://www.ebi.ac.uk/ena). After QC, 56,435 genes and 31,499 samples covering a wide range of human cell types and tissues remained. We performed a PCA on this dataset and selected 165 components representing 50% of the variation that offered the best prediction of gene function. We then selected the protein-coding genes and centred and scaled the eigenvectors for these 165 components (mean = 0, s.d. = 1) such that each component was given equal weight. The first components mostly describe tissue differences, so this normalization ensures that tissue-specific patterns do not disproportionately drive the co-regulation matrix. The co-regulation matrix is defined as the Pearson correlation between the genes from the scaled eigenvector matrix. The diagonal of the co-regulation matrix was set to zero to avoid the correlation with itself having a disproportionate effect on the association to the GWAS gene z-scores. Finally, we converted the Pearson r to z-scores.

To identify pathway and disease enrichments, we used the following databases: Human Phenotype Ontology (HPO), Kyoto Encyclopaedia of Genes and Genomes (KEGG), Reactome and Gene Ontology (GO) Biological Process, Cellular Component and Molecular Function. We have previously predicted how much each gene contributes to these gene sets, resulting in a z-score per pathway or term per gene. We collapsed genes into meta-genes in parallel with the GWAS step, to ensure compatibility with the GWAS gene z-score following the same procedure as in the GWAS pre-processing. Meta-gene z-scores were calculated as the z-score sum divided by the square root of the number of genes. Finally, all pathway z-scores were scaled (mean = 0, s.d. = 1).

For each GWAS, both real and simulated, we carried out the rank-based inverse normal transformation of GWAS z-scores to ensure that outliers would not have disproportionate weights. Owing to limitations in the PASCAL methodology that result in ties at a minimum significance level of 1*x*10^12^ for highly significant genes, we used the minimum SNP p-value from the GWAS to identify the most significant gene and resolve the tie. We then used a linear model to correct for gene length, as longer genes will typically harbour more SNPs. Sometimes, two (or more) genes will be so close to one another that their GWAS gene z-scores are highly correlated, violating the assumptions of the linear model. Thus, genes with a Pearson correlation *r* ≥ 0.8 in the 10,000 GWAS permutations were collapsed into ‘meta-genes’ and treated as one gene. Meta-gene z-scores were averaged across the input z-scores. Lastly, the GWAS z-scores of the meta-genes were scaled (mean = 0, s.d. = 1). We used a GLS regression to associate the GWAS gene z-scores to the pathway z-scores and co-regulation z-scores (described below). These two analyses result in pathway enrichments and core gene prioritizations, respectively. We used the gene–gene correlation matrix derived from the 10,000 permutations as a measure of conditional covariance of the error term (*ω*) in the GLS to account for the relationships between genes due to LD and proximity. The pseudo-inverse of *ω* is used as a substitute for *ω*^−1^.

The formula of the GLS is *β* = (*X*^*T*^ *ω*^−1^*X*)^−1^*X*^*T*^ *ω*^−1^*y*, where *β* is the estimated effect size of a pathway, term or gene from the co-regulation matrix, *ω* is the gene–gene correlation matrix, X is the design matrix of real GWAS z-scores and y is the vector of gene z-scores per pathway, term or gene from the co-regulation matrix. As we standardized the predictors, we did not include an intercept in the design matrix and X contains only one column with the real GWAS gene z-scores. We estimated *β* for the 10,000 random GWASs in the same way and subsequently used them to estimate the empirical p-value for *β*. We present full results for each age in Tables S3 to S10.

### Genetic correlations

We used LD Score regression to calculate genetic correlations among the test statistics generated using Equation 5 at ages 41, 43, 45, 47, 49, 51, 53, 55, and among these ages and other phenotypes using publicly available GWAS summary data. We present these estimates in Figure 4.

### Mendelian Randomisation

We calculated the causal effect estimates that ANM at different ages has on various traits using Mendelian Randomization (MR); a statistical method which utilizes the randomized inheritance of genetic variations in the population to estimate the potential causal effect a modifiable risk factor or exposure has on a health-related outcome of interest [23, 24]. The genetic variants used as instrumental variables (IVs) for our exposure were selected to have a genome-wide significant association with the exposure (*p <* 5 · 10^−8^) and were then pruned using linkage disequilibrium (LD) distance to ensure that they were independent. This was done using the ‘ld_clump’ function of the ‘ieugwasr’ R package [25] with default settings (clump_kb = 10000, clump_r2 = 0.001, clump_p = 0.99, pop = “EUR”). After the IVs of our exposure were selected, their association effects were then obtained for each of our outcome traits of interest. A single-sided t-test was carried out to check if the IVs had a stronger association with the outcome than with the exposure and were subsequently removed if so (for violating the MR assumptions). The two sets of association effects were then harmonized and used to calculate the causal effect estimates using the Inverse Variance Weighted method found in the ‘TwoSampleMR’ R package [26]. This analysis was repeated for each varying age of our exposure. It is important to note that in the case of Educational attainment as an outcome, there were little exposure IVs that overlapped with the outcome genetic variants, especially as the age increased, hence in the presence of a single IV, a Wald ratio was used to calculate the MR causal effect estimate. Moreover, when the trait was of a case-control nature, the effective sample size was calculated using the following formula: (4*cases*controls)/(cases+controls) [27].

## Data Availability

The BayesW model was executed with the software Hydra, with full open source code available at https://github.com/medical-genomics-group/hydra [28]. The scripts used to execute CAMP model are available at https://github.com/svenojavee/CAMP. Age-specific summary statistic estimates are released publicly on Dryad: https://doi.org/10.5061/dryad.nvx0k6dx5.

## Author contributions

SEO and MRR conceived and designed the study. SEO and ZK conceived and derived the significance testing. SEO, LD, MP and MRR conducted the analysis. KL, KF, RM provided study oversight and contributed data. SEO and MRR wrote the paper. All authors approved the final manuscript prior to submission.

## Data availability

This project uses UK Biobank data under project 35520. UK Biobank genotypic and phenotypic data is available through a formal request at (http://www.ukbiobank.ac.uk). The UK Biobank has ethics approval from the North West Multi-centre Research Ethics Committee (MREC). For access to be granted to the Estonian Biobank genotypic and corresponding phenotypic data, a preliminary application must be presented to the oversight committee, who must first approve the project, ethics permission must then be obtained from the Estonian Committee on Bioethics and Human Research, and finally a full project must be submitted and approved by the Estonian Biobank. This project was granted ethics approval by the Estonian Committee on Bioethics and Human Research (https://genomics.ut.ee/en/biobank.ee/data-access). Age-specific summary statistic estimates are released publicly on Dryad: https://doi.org/10.5061/dryad.nvx0k6dx5.

## Code availability

The BayesW model was executed with the software Hydra, with full open source code available at https://github.com/medical-genomics-group/hydra [28]. The scripts used to execute CAMP model is available at https://github.com/svenojavee/CAMP. R version 4.2.1 is available at https://www.r-project.org/.

## Acknowledgements

This project was funded by an SNSF Eccellenza Grant to MRR (PCEGP3-181181), and by core funding from the Institute of Science and Technology Austria. K.L. and R.M. were supported by the Estonian Research Council grant 1911. Estonian Biobank computations were performed in the High Performance Computing Centre, University of Tartu. We thank Triin Laisk for her valuable insights and comments that helped greatly. We would like to acknowledge the participants and investigators of UK Biobank and Estonian Biobank studies. This project uses UK Biobank data under project number 35520.

## Supplementary Tables

**Table S1.** Regions with a genome-wide significant age-specific effect on age-at-menopause not replicated in the Estonian Biobank. Although these SNPs did not have a significant slope in the replication data set, the genome-wide significant non-zero slopes indicate that the constant effect assumption might be too stringent when analysing the effect of these variants. For each SNP we tested the significance of the slope parameter using the CAMP model. The results were then LD clumped such that the index SNPs would have a p-value below 5 · 10^−8^ and SNPs could be added to a clump if they were 1Mb from the index SNP, they were correlated with *r*^2^ *>* 0.05 and they were nominally significant (*p <* 0.05). We then used the COJO method from the GCTA software (see Methods) to find clumps with independent signals by conducting stepwise selection of index SNPs in 1Mb window and we considered SNPs independent if they had a p-value below 5 · 10^−8^ in the joint model. The candidates for significant slope were replicated in the Estonian Biobank. Replication was defined as p-value being lower than 0.05 and the direction of the effect size same in both the original analysis and the replication analysis. The effect size estimates are reported on the log hazard scale. The column Nearest gene is mapped from the SNP using ANNOVAR software, * in that column denotes intergenic regions; chromosome X nearest gene was determined by using the UCSC Genome Browser. The column Strongest evidence p-value indicates the p-value at age when there is strongest evidence for an effect.

| SNP | Chr | Position | Eff/Oth | MAF | Nearest gene | Yearly effect change | Slope p-value | Strongest evidence p-value |
| --- | --- | --- | --- | --- | --- | --- | --- | --- |
| rs115441717 | 1 | 17,587,645 | A/G | 0.005 | PADI3 | -0.0036 | $5.37 \times 10^{-10}$ | $7.51 \times 10^{-10}$ |
| rs186475193 | 1 | 66,520,884 | A/T | 0.003 | PDE4B | -0.0036 | $8.05 \times 10^{-11}$ | $1.30 \times 10^{-10}$ |
| rs61813814 | 1 | 178,259,880 | A/G | 0.004 | RASAL2 | -0.0035 | $3.34 \times 10^{-9}$ | $3.18 \times 10^{-7}$ |
| rs9662346 | 1 | 246,893,259 | G/A | 0.271 | SCCPDH | 0.0039 | $5.59 \times 10^{-9}$ | $1.20 \times 10^{-3}$ |
| rs1406295 | 2 | 27,689,700 | G/A | 0.393 | IFT172 | -0.0038 | $5.21 \times 10^{-9}$ | $1.92 \times 10^{-50}$ |
| rs2293269 | 2 | 48,844,968 | C/G | 0.352 | GTF2A1L* | -0.0044 | $3.30 \times 10^{-11}$ | $1.79 \times 10^{-20}$ |
| rs117292624 | 3 | 5,813,506 | A/T | 0.006 | AC027119.1* | -0.0034 | $6.36 \times 10^{-9}$ | $3.66 \times 10^{-8}$ |
| rs1317571 | 3 | 49,910,657 | T/G | 0.177 | ACTBP13* | -0.0042 | $2.99 \times 10^{-10}$ | $5.27 \times 10^{-12}$ |
| rs9818740 | 3 | 135,939,586 | A/G | 0.274 | RP11-463H24.1* | -0.0042 | $1.35 \times 10^{-10}$ | $1.01 \times 10^{-14}$ |
| rs149660889 | 4 | 7,769,181 | C/T | 0.006 | AFAP1* | -0.0035 | $1.61 \times 10^{-9}$ | $3.36 \times 10^{-7}$ |
| rs7661090 | 4 | 13,571,901 | T/C | 0.105 | BOD1L1 | -0.0040 | $3.45 \times 10^{-10}$ | $1.42 \times 10^{-18}$ |
| rs11099599 | 4 | 84,367,050 | A/G | 0.492 | HELQ | 0.0048 | $3.28 \times 10^{-13}$ | $6.18 \times 10^{-130}$ |
| rs116822956 | 5 | 6,790,631 | G/T | 0.024 | RP11-332J15.1* | -0.0042 | $9.96 \times 10^{-12}$ | $3.42 \times 10^{-3}$ |
| rs11737992 | 5 | 175,953,121 | T/C | 0.309 | RNF44* | -0.0042 | $1.88 \times 10^{-10}$ | $1.58 \times 10^{-30}$ |
| rs76706863 | 6 | 28,291,052 | G/T | 0.018 | ZSCAN31* | -0.0036 | $3.73 \times 10^{-9}$ | $1.27 \times 10^{-6}$ |
| rs138131857 | 6 | 29,654,098 | A/G | 0.018 | ZFP57* | -0.0041 | $2.80 \times 10^{-11}$ | $1.66 \times 10^{-10}$ |
| rs11752373 | 6 | 90,583,601 | A/T | 0.045 | CASP8AP2 | -0.0037 | $3.79 \times 10^{-9}$ | $3.22 \times 10^{-6}$ |
| rs140020429 | 6 | 109,831,159 | T/C | 0.006 | AK9 | -0.0034 | $4.88 \times 10^{-9}$ | $2.77 \times 10^{-6}$ |
| rs117109951 | 7 | 123,057,448 | C/T | 0.003 | IQUB | -0.0034 | $6.25 \times 10^{-9}$ | $1.06 \times 10^{-6}$ |
| rs10093345 | 8 | 37,872,776 | C/T | 0.259 | EIF4EBP1* | -0.0060 | $2.33 \times 10^{-18}$ | $3.02 \times 10^{-98}$ |
| rs76637775 | 8 | 69,027,721 | G/A | 0.008 | PREX2 | -0.0036 | $4.68 \times 10^{-9}$ | $7.68 \times 10^{-7}$ |
| rs138790299 | 8 | 80,383,022 | G/A | 0.003 | RP11-758H6.1* | -0.0035 | $6.67 \times 10^{-10}$ | $3.18 \times 10^{-7}$ |
| rs112136293 | 8 | 135,941,782 | A/T | 0.003 | RP11-1057B8.2* | -0.0035 | $1.11 \times 10^{-9}$ | $1.24 \times 10^{-6}$ |
| rs117273292 | 9 | 91,945,999 | C/T | 0.03 | SECISBP2 | -0.0037 | $1.72 \times 10^{-9}$ | $9.40 \times 10^{-7}$ |
| rs113357099 | 10 | 75,065,684 | A/G | 0.083 | TTC18 | 0.0041 | $3.57 \times 10^{-9}$ | $8.83 \times 10^{-4}$ |
| rs79293542 | 10 | 97,809,854 | C/T | 0.034 | CCNJ* | -0.0042 | $1.02 \times 10^{-11}$ | $1.80 \times 10^{-26}$ |
| rs11245450 | 10 | 126,658,075 | A/G | 0.420 | ZRANB1 | -0.0039 | $3.67 \times 10^{-9}$ | $9.68 \times 10^{-32}$ |
| rs655139 | 11 | 102,269,679 | G/T | 0.082 | TMEM123 | -0.0038 | $4.45 \times 10^{-9}$ | $2.49 \times 10^{-2}$ |
| rs3782232 | 12 | 57,116,249 | A/G | 0.071 | NACA | -0.0041 | $1.64 \times 10^{-10}$ | $5.04 \times 10^{-64}$ |
| rs80256675 | 12 | 68,088,664 | C/A | 0.007 | RP11-43N5.1* | -0.0036 | $2.10 \times 10^{-9}$ | $2.86 \times 10^{-6}$ |
| rs11180609 | 12 | 76,040,392 | C/T | 0.063 | RP11-114H23.1 | -0.0040 | $4.60 \times 10^{-10}$ | $7.93 \times 10^{-12}$ |
| rs145313906 | 12 | 83,821,558 | A/T | 0.003 | RP11-384P14.1* | -0.0038 | $7.07 \times 10^{-12}$ | $5.35 \times 10^{-12}$ |
| rs2954111 | 12 | 122,134,415 | C/T | 0.36 | TMEM120B* | -0.0048 | $1.45 \times 10^{-13}$ | $2.45 \times 10^{-17}$ |
| rs7322160 | 13 | 61,061,456 | C/T | 0.328 | TDRD3 | 0.0045 | $1.81 \times 10^{-11}$ | $1.89 \times 10^{-42}$ |
| rs146576015 | 16 | 60,155,191 | G/A | 0.010 | RP11-430C1.1* | -0.0035 | $6.00 \times 10^{-9}$ | $4.52 \times 10^{-8}$ |
| rs140438673 | 16 | 85,217,665 | A/G | 0.007 | CTC-786C10.1 | -0.0035 | $6.05 \times 10^{-9}$ | $2.29 \times 10^{-8}$ |
| rs13331995 | 16 | 89,912,063 | C/T | 0.346 | SPIRE2 | -0.0045 | $7.59 \times 10^{-12}$ | $2.90 \times 10^{-29}$ |
| rs62097189 | 18 | 32,191,404 | G/A | 0.003 | DTNA | -0.0034 | $2.22 \times 10^{-9}$ | $1.13 \times 10^{-12}$ |
| rs148228698 | 19 | 2,417,585 | A/G | 0.008 | TMPRSS9 | -0.0035 | $6.61 \times 10^{-9}$ | $2.50 \times 10^{-7}$ |
| rs7253685 | 19 | 55,845,386 | G/A | 0.378 | TMEM150B | -0.0067 | $2.25 \times 10^{-24}$ | $1.57 \times 10^{-70}$ |
| rs16991615 | 20 | 5,948,227 | A/G | 0.066 | MCM8 | 0.0091 | $1.13 \times 10^{-36}$ | $1.13 \times 10^{-36}$ |
| rs5951636 | X | 21,841,633 | C/T | 0.377 | MBTPS2* | -0.0041 | $3.11 \times 10^{-10}$ | $3.69 \times 10^{-18}$ |
| rs12559662 | X | 44,155,570 | T/C | 0.101 | EFHC2 | -0.0038 | $3.69 \times 10^{-9}$ | $1.06 \times 10^{-14}$ |

## Supplementary Figures

**Figure S1.**
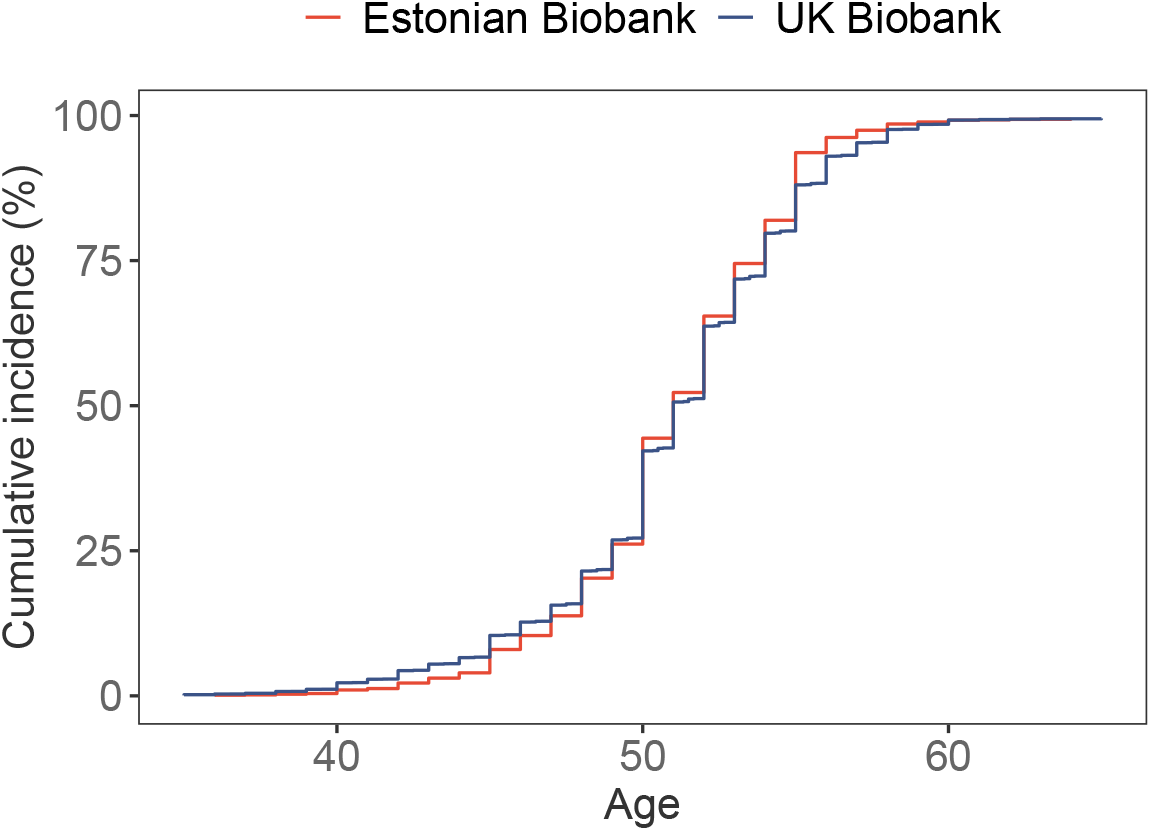
Cumulative incidence curves of menopause. Cumulative incidence curves take into account the competing risk of death. The distributions of age-at-menopause in the UK and Estonian Biobanks are relatively similar, although more menopauses happen somewhat earlier in UK Biobank than in the Estonian Biobank.

**Figure S2.**
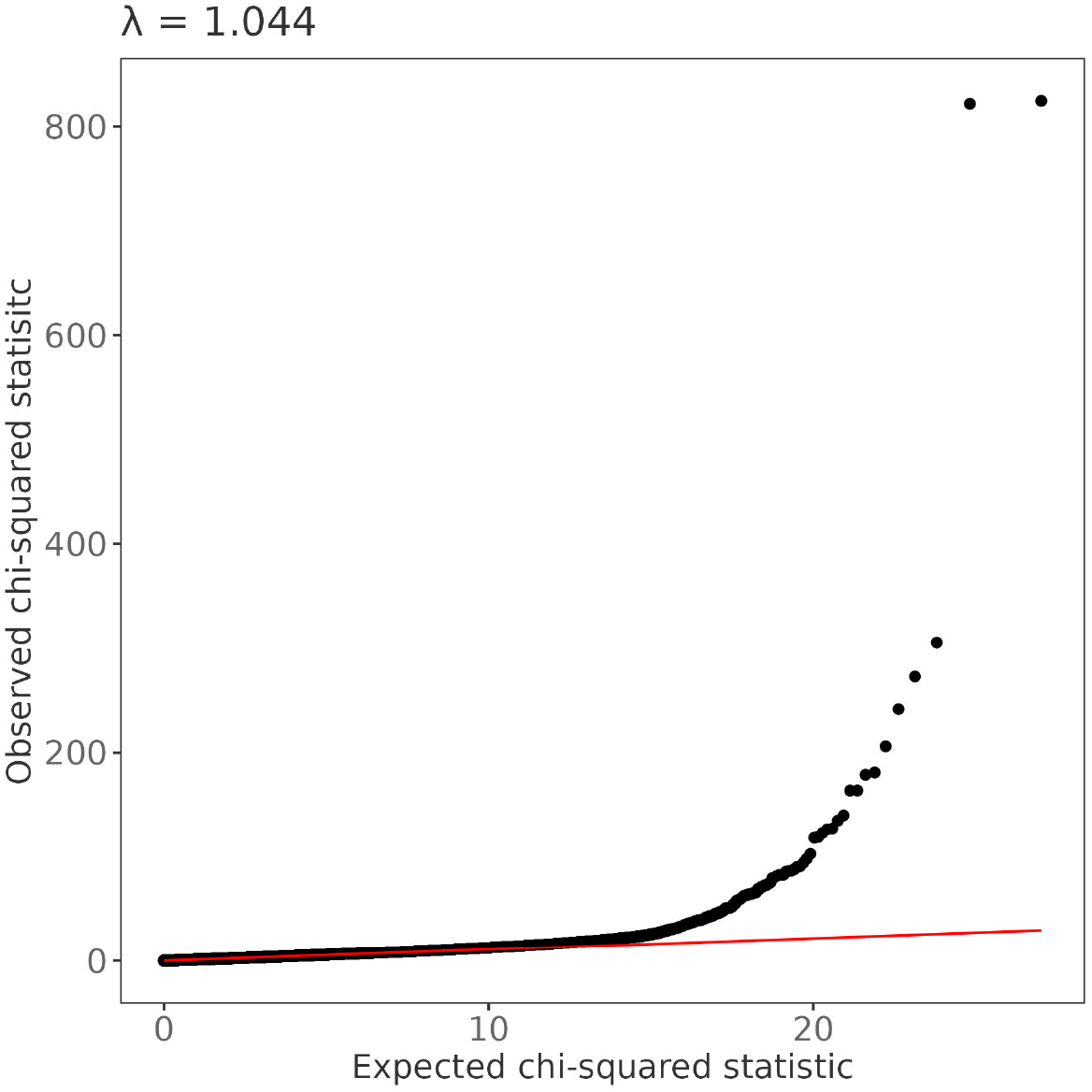
QQ plot of age-at-menopause GWAS using CAMP model. The x-axis uses the 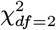 distribution quantiles as under the null hypothesis of no effect (*β*^0^ = 0 and *β*^1^ = 0) the test statistic follows that distribution. The genomic inflation factor is 1.044, median of 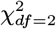 distribution is 1.386.

**Figure S3.**
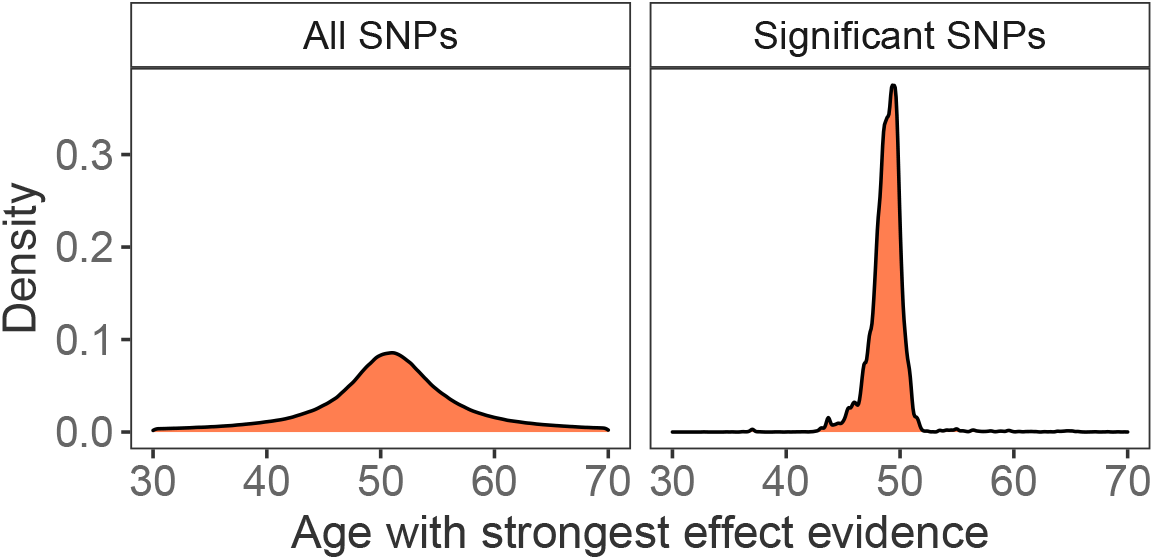
Distribution of ages with the most evidence for an effect size. Distribution of the ages when the maximum chi-squared statistic is achieved (calculated using Equation 10) over all SNPs and significant SNPs only. Even though the distributions have similar centres (median age of 51 and 49 for all SNPs and significant SNPs, respectively), the standard deviation of all SNP age distribution is 3.7 times higher than significant SNP age distribution.

**Figure S4.**
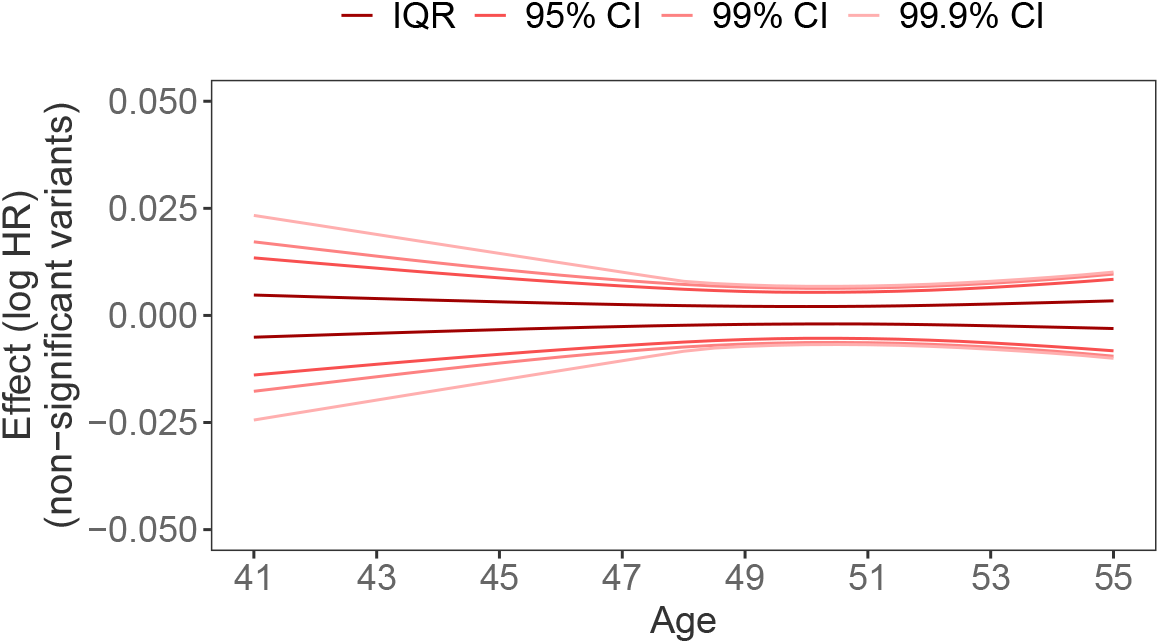
Effect sizes (log HR) for nominally non-significant effects by summarising the effects between the interquartile range, 95% CI, 99% CI or 99.9% CI. We observe that compared to the significant effect sizes, the non-significant effects are much smaller and do not vary much across ages. Nominally insignificant effects are defined as *p >* 0.05.

**Figure S5.**
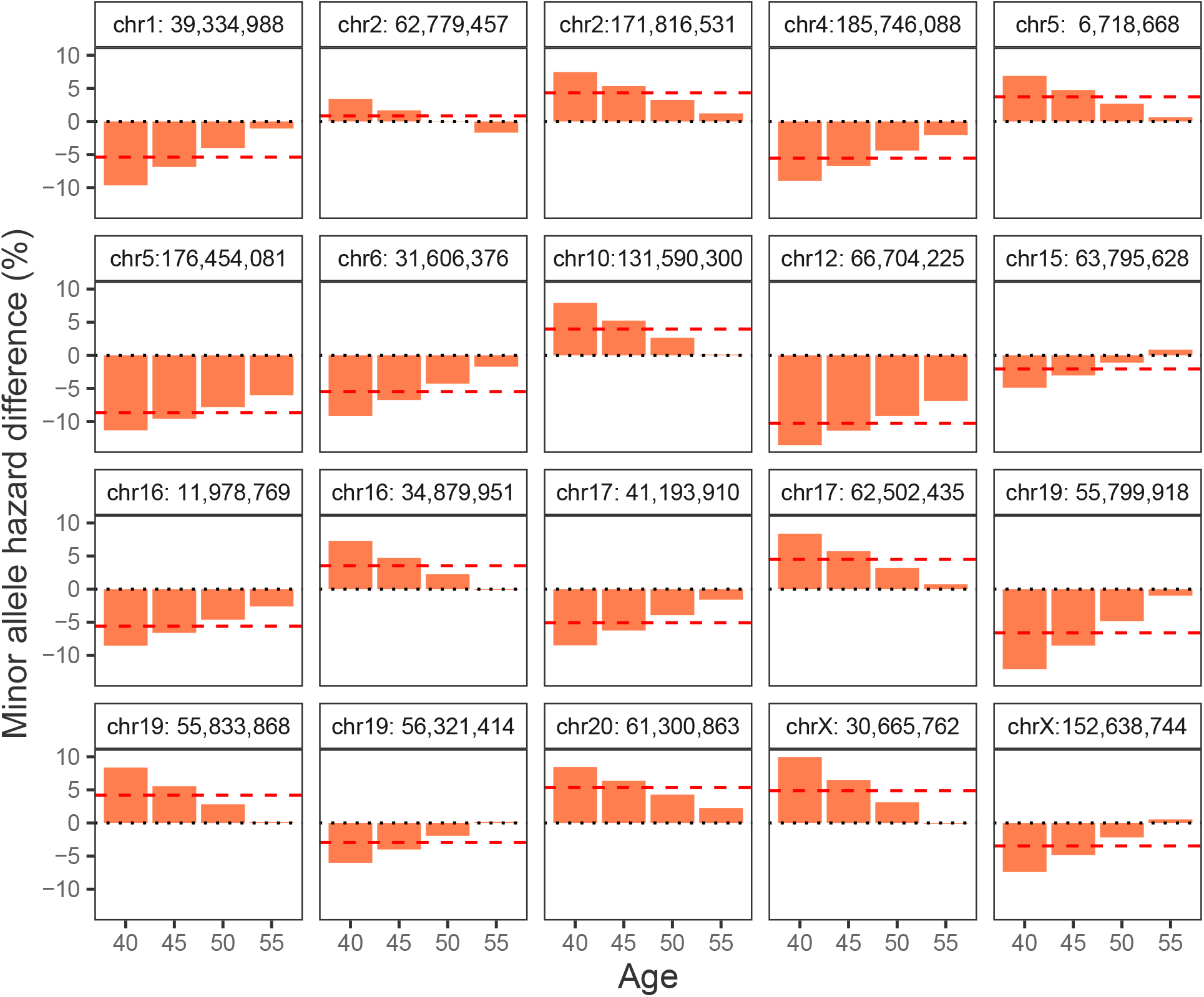
Change in hazard for the variants with a significant age-specific effects. Illustration of the effect size change for the variants with an age-specific effects in Table 2.

**Figure S6.**
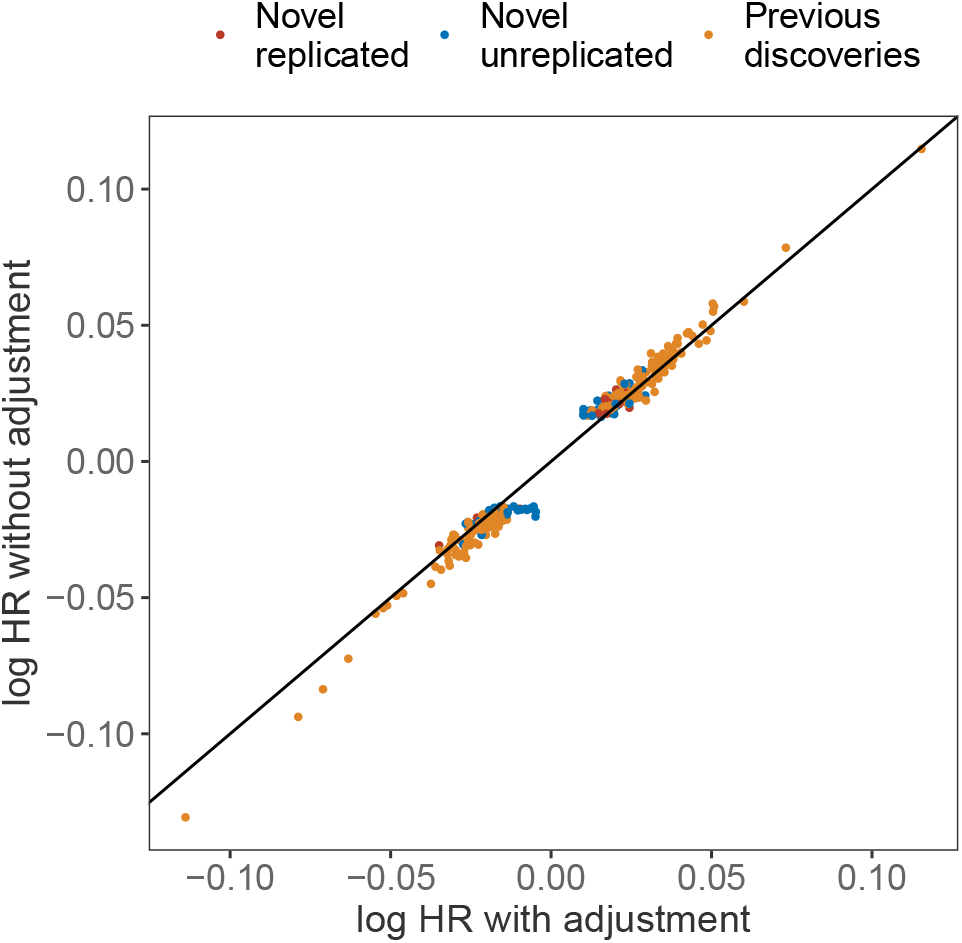
Sensitivity analysis for a model with and without adjusting hormone replacement therapy (HRT) as a time-varying covariate. Log hazard ratio effect size estimates for the 312 age-at-natural menopause (ANM) associations with (x-axis) and without (y-axis) HRT as covariate. The effect sizes shown are the age between 45-52 where the maximum *χ*^2^ statistic value was obtained for either model.

## References

1. Ojavee, S. E. et al. Genomic architecture and prediction of censored time-to-event phenotypes with a bayesian genome-wide analysis. Nature Communications 12, 2337 (2021).

2. Pedersen, E. M. et al. Accounting for age of onset and family history improves power in genome-wide association studies. The American Journal of Human Genetics 109, 417–432 (2022).

3. Shin, J. et al. Age at menopause and risk of heart failure and atrial fibrillation: a nationwide cohort study. European Heart Journal 43, 4148–4157 (2022).

4. Nash, Z., Al-Wattar, B. H. & Davies, M. Bone and heart health in menopause. Best Practice & Research Clinical Obstetrics & Gynaecology (2022).

5. Collaborative Group on Hormonal Factors in Breast Cancer and others. Menarche, menopause, and breast cancer risk: individual participant meta-analysis, including 118 964 women with breast cancer from 117 epidemiological studies. The Lancet Oncology 13, 1141–1151 (2012).

6. Murabito, J. M., Yang, Q., Fox, C., Wilson, P. W. & Cupples, L. A. Heritability of age at natural menopause in the framingham heart study. The Journal of Clinical Endocrinology & Metabolism 90, 3427–3430 (2005).

7. Stolk, L. et al. Meta-analyses identify 13 loci associated with age at menopause and highlight dna repair and immune pathways. Nature genetics 44, 260–268 (2012).

8. Day, F. R. et al. Large-scale genomic analyses link reproductive aging to hypothalamic signaling, breast cancer susceptibility and brca1-mediated dna repair. Nature genetics 47, 1294–1303 (2015).

9. Ruth, K. S. et al. Genetic insights into biological mechanisms governing human ovarian ageing. Nature 596, 393–397 (2021).

10. Jiang, X., Holmes, C. & McVean, G. The impact of age on genetic risk for common diseases. PLoS genetics 17, e1009723 (2021).

11. Winkler, T. W. et al. The influence of age and sex on genetic associations with adult body size and shape: a large-scale genome-wide interaction study. PLoS genetics 11, e1005378 (2015).

12. Levine, M. E. et al. Menopause accelerates biological aging. Proceedings of the National Academy of Sciences 113, 9327–9332 (2016).

13. Ward, L. D. et al. Rare coding variants in dna damage repair genes associated with timing of natural menopause. Human Genetics and Genomics Advances 3, 100079 (2022). URL https://www.sciencedirect.com/science/article/pii/S2666247721000609.

14. Robinson, M. R. et al. Genotype–covariate interaction effects and the heritability of adult body mass index. Nature genetics 49, 1174–1181 (2017).

15. Joshi, P. K. et al. Variants near chrna3/5 and apoe have age-and sex-related effects on human lifespan. Nature communications 7, 1–7 (2016).

16. Patxot, M. et al. Probabilistic inference of the genetic architecture underlying functional enrichment of complex traits. Nature Communications 12, 6972 (2021).

17. Tasa, T. et al. Genetic variation in the estonian population: pharmacogenomics study of adverse drug effects using electronic health records. European Journal of Human Genetics 27, 442–454 (2019).

18. Purcell, S. et al. Plink: a tool set for whole-genome association and population-based linkage analyses. The American journal of human genetics 81, 559–575 (2007).

19. Yang, J. et al. Conditional and joint multiple-snp analysis of gwas summary statistics identifies additional variants influencing complex traits. Nature genetics 44, 369–375 (2012).

20. Yang, J., Lee, S. H., Goddard, M. E. & Visscher, P. M. GCTA: a tool for genome-wide complex trait analysis, journal=American journal of human genetics 88, 76–82 (2011). PMC3014363[pmcid].

21. Staley, J. R. et al. PhenoScanner: a database of human genotype–phenotype associations. Bioinformatics 32, 3207–3209 (2016).

22. Kamat, M. A. et al. PhenoScanner v2: an expanded tool for searching human geno-type–phenotype associations. Bioinformatics 35, 4851–4853 (2019).

23. Sanderson, E. et al. Mendelian randomization. Nature Reviews Methods Primers 2, 6 (2022).

24. Davey Smith, G. & Hemani, G. Mendelian randomization: genetic anchors for causal inference in epidemiological studies. Human molecular genetics 23, R89–R98 (2014).

25. Hemani, G. ieugwasr: R interface to the ieu gwas database api. R package version 0.1 5 (2020).

26. Hemani, G. et al. The mr-base platform supports systematic causal inference across the human phenome. eLife 7, e34408 (2018). URL https://elifesciences.org/articles/34408.

27. Han, B. & Eskin, E. Random-effects model aimed at discovering associations in meta-analysis of genome-wide association studies. The American Journal of Human Genetics 88, 586–598 (2011).

28. Robinson, M. hydra (version v1.0). Zenodo (2021). URL http://doi.org/10.5281/zenodo.4555238.

